# Seasonality of acute kidney injury phenotypes in England: an unsupervised machine learning classification study of electronic health records

**DOI:** 10.1101/2023.03.17.23287400

**Authors:** Hikaru Bolt, Anne Suffel, Julian Matthewman, Frank Sandmann, Laurie Tomlinson, Rosalind Eggo

## Abstract

**Background:** Acute Kidney Injury (AKI) is a multifactorial condition which presents a substantial burden to healthcare systems. There is limited evidence on whether it is seasonal. We sought to investigate the seasonality of AKI hospitalisations in England and use unsupervised machine learning to explore clustering of underlying comorbidities, to gain insights for future intervention.

**Methods:** We used Hospital Episodes Statistics linked to the Clinical Practice Research Datalink to describe the overall incidence of AKI admissions between 2015-2019 weekly by demographic and admission characteristics. We carried out dimension reduction on 850 diagnosis codes using multiple correspondence analysis and applied k-means clustering to classify patients. We phenotype each group based on the dominant characteristics and describe the seasonality of AKI admissions by these different phenotypes.

**Findings:** Between 2015-2019, weekly AKI admissions peaked in winter, with additional summer peaks related to periods of extreme heat. Winter seasonality was more evident in those diagnosed with AKI on admission. From the cluster classification we describe six phenotypes of people admitted to hospital with AKI. Among these, seasonality of AKI admissions was observed among people who we described as having a multimorbid phenotype, established risk factor phenotype, and general AKI phenotype.

**Interpretation:** We demonstrate winter seasonality of AKI admissions in England, particularly among those with AKI diagnosed on admission, suggestive of community triggers. Differences in seasonality between phenotypes suggests some groups may be more likely to develop AKI as a result of these factors. This may be driven by underlying comorbidity profiles or reflect differences in uptake of seasonal interventions such as vaccines.

**Funding:** This study was funded by the National Institute for Health and Care Research (NIHR) Health Protection Research Unit (HPRU) in Modelling and Health Economics, a partnership between UK Health Security Agency (UKHSA), Imperial College London, and London School of Hygiene and Tropical Medicine. The views expressed are those of the authors and not necessarily those of the National Health Service, NIHR, UK Department of Health or UKHSA.

**Research in context:** *Evidence before this study:* We searched for articles in Medline using the terms (“Seasons/” OR “Seasons”) AND (“Acute Kidney Injury/” OR “Acute Kidney Injury” OR “AKI” OR “ARF”). We also search Embase using the terms (“Seasonal variation/” OR “Seasonal variation” OR “Season/” OR “Season”) AND (“Acute kidney failure/” OR “Acute kidney failure” OR “AKI” OR “ARF”. Articles published until 20/01/2023 in any language were included. Only two studies investigated seasonality of AKI in the UK and indicated winter increases in admissions. However, both studies aggregate AKI hospitalisations into quarterly counts and therefore were unable to show acute weekly changes in AKI admissions and timings of peaks. Studies outside of the UK varied in their conclusions of summer or winter increases in AKI admissions and the profile of patients driving this variation.

*Added value of this study:* This is the largest and most granular investigation of AKI seasonality in England, investigating 198,754 admissions in a weekly time series detecting acute changes in incidence and differences in peaks year to year. We demonstrate consistent peaks in the winter as well as acute peaks in the summer. Most records indicated AKI was diagnosed on admission therefore suggestive of community triggers of AKI. We included more data on the profile of patients than previously published studies. Our novel approach to investigate the profile of seasonal admissions using unsupervised machine learning suggests some groups may be more affected by seasonal triggers than others.

*Implications of all the available evidence:* AKI is a common syndrome which leads to hospitalisation with a significant burden on the health system. We demonstrate a conclusive seasonal pattern to AKI admissions which has important implications on healthcare provision planning, public health, and clinical practice in England. Future research on AKI should take into account seasonality; uncertainty remains on the main drivers and aetiology of the seasonal patterns observed.

## Background

Acute kidney injury (AKI) is a syndrome defined by rapid decline in kidney function from hours to days leading to disruption in metabolic, electrolyte, and fluid homeostasis (1). Between 20-25% of hospitalised adults have AKI, and it is associated with longer duration of stay and a 4-16 fold increase in odds of death following hospitalisation (1–3). The heterogeneity of the condition and its triggers and the wide range of risk factors makes it difficult to identify important mechanisms which can be modified to reduce the incidence of AKI (1).

Previous studies have demonstrated a seasonal winter pattern to AKI hospital admissions (4–6). Data from a Welsh automated electronic AKI reporting system found an increase in AKI alerts during winter in primary and secondary care (4). Furthermore, a study in Japan indicated an increased odds of AKI in winter months with seasonality most pronounced for patients primarily diagnosed with cardiovascular and pulmonary admission codes, and when AKI was diagnosed on the day of admission (5). While winter increases in AKI suggest association with infections (7), other conditions associated with AKI such as heart failure and myocardial infarction also have seasonal patterns (8–11).

Given the high incidence and complex, multifactorial aetiology of AKI the condition is well suited to analysis using machine learning (ML) (12–14). ML is increasingly used to analyse electronic health records (EHR) for risk prediction models, causal inference, text mining, and phenotypic discovery methods (12–14). Previous studies using unsupervised clustering classification of EHR data include studies such as identifying clinical phenotypes of heart failure, Alzheimer’s disease, and chronic obstructive pulmonary disease to describe the diversity of expression, progression, and aetiology of patients experiencing the same disease (14–16). The primary benefit of unsupervised clustering classification is the ability to analyse large datasets without pre-specifying hypotheses or interactions, and without limiting the number of features included to phenotype patients (17). ML methods could uncover new and important phenotypes of AKI not previously considered for detailed epidemiological investigation, and new targets for intervention.

Therefore, in this study using routine primary and secondary care data from England, we sought to firstly determine whether there is seasonality in AKI admissions in England, and any associations with age and gender, and secondly to use unsupervised ML clustering approaches to investigate AKI phenotypes, and whether these also demonstrated seasonality.

## Methods

### Data source

We used linked primary and secondary care data from England in CPRD GOLD, which is a large primary care database collecting longitudinal EHRs from participating GPs representing 21 million patients with 3 million currently registered (18). Data is quality assured and includes demographic characteristics, diagnoses and symptoms, drug exposures, vaccination history, laboratory tests, and referrals to secondary care (18). Data are recorded using Read codes, a standardised hierarchical coding structure to describe a patient’s consultation and condition.

CPRD has been shown to be representative of the UK population by age, sex, and ethnicity (17).

In 2019, 52% of CPRD GOLD patients were linked to hospital episode statistics (HES) which records hospital admissions, attendances to Accident & Emergency, and outpatient appointments to all NHS hospitals. Data in HES are recorded using the International Classification of Diseases version 10 (ICD-10) codes, where each code represents a diagnosis, which are grouped under 22 headings in a hierarchical structure.

### Study population

We defined the source population as all patients recorded between January 2015 – December 2019, that met research acceptable quality control standards (18). We defined the cohort as patients admitted to hospital with an AKI ICD-10 code (ICD-10 N-17 and N-19) in any diagnostic position during an admission (Supplementary table 1).

### Feature selection

We extracted linked primary care records for the study population which were stored as Read codes. We mapped the Read codes to the relevant hierarchy from specific to general terms, and we prepared the features for clustering at level 3 (e.g. G30.. - Acute myocardial infarction).

We included diagnosis codes as features for the cluster classification (Supplementary table 2), and age and sex were included as supplementary variables, used to describe the cluster but not included in the cluster classification algorithm. We excluded codes relating to symptoms, medical procedures, and lifestyle factors (Supplementary table 3), as well as Read code chapter Z (Unspecified conditions) to reduce the number of features included to improve processing capacity. We excluded features recorded less than 100 times in the observation period across all patients in order to reduce the computational burden, and made the assumption that these features will not have a material impact on clusters formed due to the low frequency. Diagnosis codes relating to infectious diseases (Chapter A - Infectious and parasitic diseases; Chapter H0 - Acute respiratory infections; Chapter H1 - Other upper respiratory tract diseases; Chapter H2 - Pneumonia and influenza; Chapter K190 - Urinary tract infection, site not specified) were removed if they were more than 30 days before or anytime after the AKI hospitalisation. This was done in order to reflect the acute nature of these diagnoses, and time bounding these codes selected the diagnoses possibly associated with subsequent development of AKI. Without this, infection codes unrelated to AKI in time would have a dominant impact in the formation of clusters.

To prepare for cluster classification, we transformed the data into a matrix indicating the presence and absence of codes for each patient.

#### Dimension reduction

We used Multiple Correspondence Analysis (MCA) as a dimension reduction technique (15,19). Dimension reduction improves the efficiency of clustering methods, while preserving the global structure and correlation between data points (19). We selected the optimum number of dimensions using a scree plot to observe the percentage of variance in each dimension (19). We applied the “elbow rule” to the plot to determine the number of dimensions to retain.

#### K-means clustering and phenotyping

We used k-means clustering to classify patients into groups. K-means clustering classifies patients into a pre-specified number of groups based on the distance from mean centre points that minimises the total within-cluster sum of squares (15). Patients with similar characteristics are therefore classified in the same clusters. We selected the optimum number of clusters using the *NbClust* package (20). This package calculates the optimum number of clusters using 30 different indices and aggregates the results for the user to make an assessment of the optimum number of clusters in the dataset of interest (20). Due to the computational burden of applying different indices, a random sample of 25,000 patients from the cohort were selected to apply the method. To ensure consistency, five random samples were taken.

Once we allocated patients to clusters we used the frequency of clinical codes in each cluster to describe the dominant characteristics of each, using Read code chapter level 2.

### Analysis

We described the overall incidence of AKI admissions between 2015-2019 and disaggregated by age, sex, diagnostic position of AKI code, and the day during the admission where AKI was recorded. We described the clusters of AKI patients by age, sex, and Read codes at chapter level 2. All Read codes were reviewed for describing the cluster, however 17 codes were selected for illustrative purposes and cover codes mostly commonly reported as well as being plausible risk factors for AKI. We labelled clusters with the description of the overall phenotype of the cluster based on the dominant characteristics observed. We then described the incidence of AKI admissions by the cluster phenotypes.

### Sensitivity analysis

We conducted sensitivity analysis of the cluster phenotypes by 1) restricting the cohort to those who had an AKI ICD10 code of N17 (i.e. excluding N19 codes) 2) restricting the cohort to those where AKI was recorded in a primary diagnostic position and separately a secondary diagnostic position, 3) restricting the cohort to those diagnosed with AKI on admission (day 0), 4) setting random seeds to test the reproducibility of the clustering method, and 5) changing the number of dimensions included for the cluster analysis following MCA to observe the impact on the cluster phenotypes.

## Results

### Incidence

Among the cohort of 133,488 individual patients recorded to have an admission with AKI in England, between 2015-2019, there were a total of 198,754 admissions.. 52% were male and the median age was 78 (IQR: 66-86). AKI incidence increased over the entire observation period from 34,539 admissions in 2015 to 42,326 admissions in 2019. We observed distinct peaks in AKI admissions in December and January of each year (Figure 1, Supplementary figure 1), as well as June-July.

**Figure 1:**
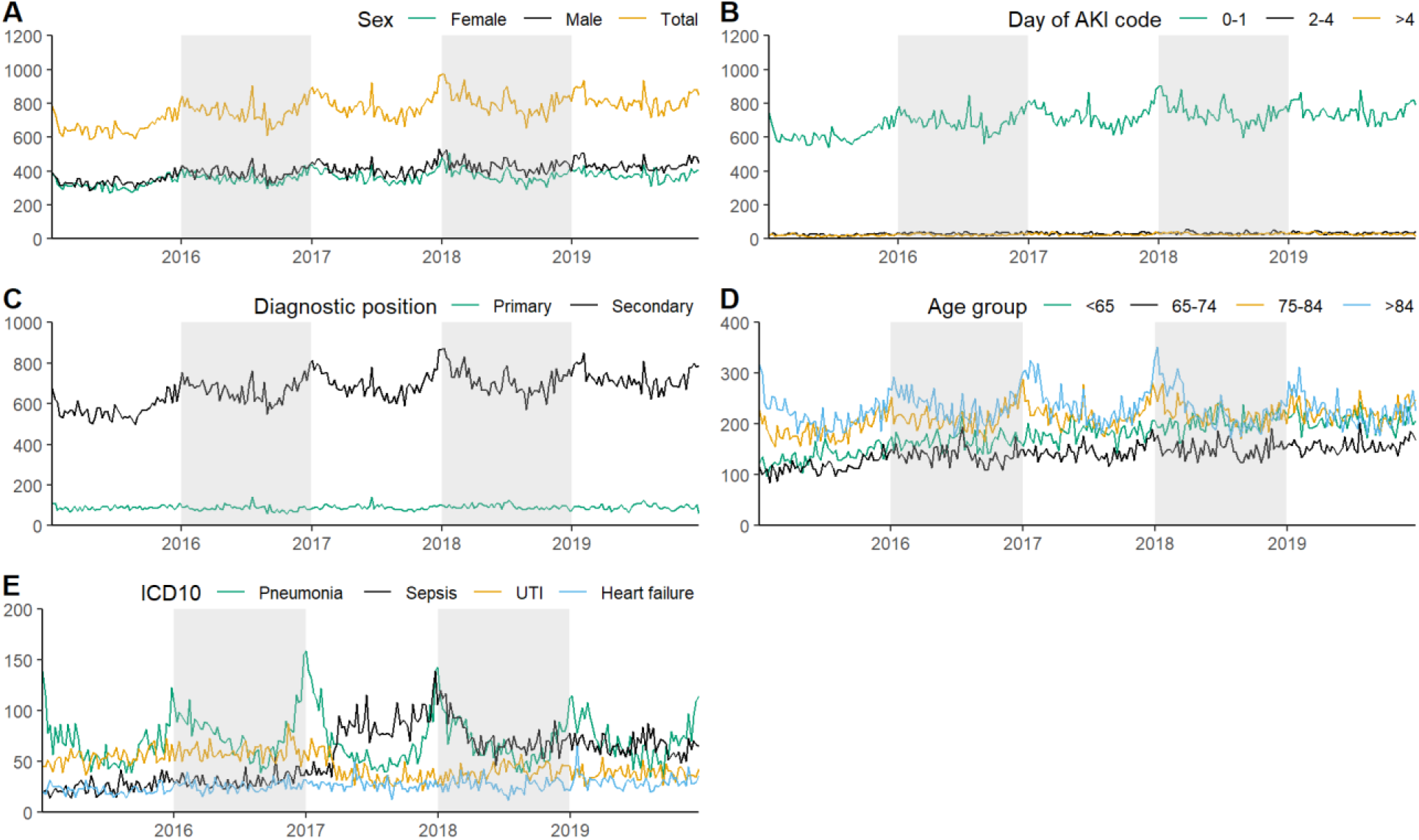
AKI admissions in HES-linked CPRD 2015-2029. Time series of weekly AKI admissions, 2015 - 2019, England total and by A) sex of patients B) day AKI code was recorded during the admission C) diagnostic position of AKI record D) age group E) primary diagnosis where AKI was a secondary code during the admission (primary diagnoses displayed make up 30% of all primary diagnoses recorded).

### Winter seasonality

There were seasonal peaks in admissions among men and women (Figure 1A), most prominently observed in people aged >75 (Figure 1D). AKI admission codes recorded on day 0-1 of the admission had evidence of seasonality, with no seasonality observed where AKI was recorded >2 days after admission (Figure 1B). Analysis of diagnostic position of AKI codes also demonstrated that seasonality is more apparent where AKI was recorded as non-primary reason for admission (diagnostic position >2) (Figure 1C). Among those with a secondary AKI code, the most common primary reasons for admissions were pneumonia, urinary tract infections (UTIs), sepsis, heart failure, and chronic obstructive pulmonary disease (COPD) (Supplementary table 4). These codes make up 30% of admissions where AKI was recorded as secondary code. Seasonality was most notable for admissions where pneumonia was the primary diagnosis (Figure 1E).

### Summer seasonality

We observed short peaks in the summer of each year; one to two weeks in duration. These peaks were observed among men and women (Figure 1A) and across age groups (Figure 1D), although not consistently across all years. Summer peaks were only observed where people were coded with AKI on day 0-1 of admission (Figure 1B), and were observed where AKI was recorded as a primary or secondary diagnostic position (Figure 1C). These periods of increased AKI admissions in the summer across all years, coincide with heatwave alerts declared by the Meteorological Office in England (21) (Supplementary figure 2).

#### Cluster classification

There were 133,488 patients that were diagnosed with AKI during an admission between 2015-2019. Among these patients there were 1,788 chapter level 3 Read codes available for dimension reduction using MCA. Of these, 938 codes were recorded less than 100 times across all patients in the time period, and were excluded. Thus 850 features were retained, which made up 99.6% of records reported among the cohort. Following exclusion of sparsely recorded variables, 130,625 patients were retained for the cluster analysis.

Following dimension reduction we retained five dimensions for cluster analysis (Supplementary figure 3). K-means clustering was applied to the five dimensions, and the analysis of different indices selected between two to 10 clusters as the optimum number of clusters (Supplementary figure 4). More indices selected two and six as the optimum number of clusters for the dataset, therefore for the analysis we presented the cluster phenotypes up to k = 6 (Supplementary figure 5).

As the number of clusters increased, further clusters were generally created as subsets of one existing cluster at each step of k (Figure 2). One exception was the creation of cluster 2 at k = 3, which was formed as a large branch from two existing clusters. At k = 4 a small cluster was formed defined by a group of patients characterised by non-specific coding (discussed further later).

**Figure 2:**
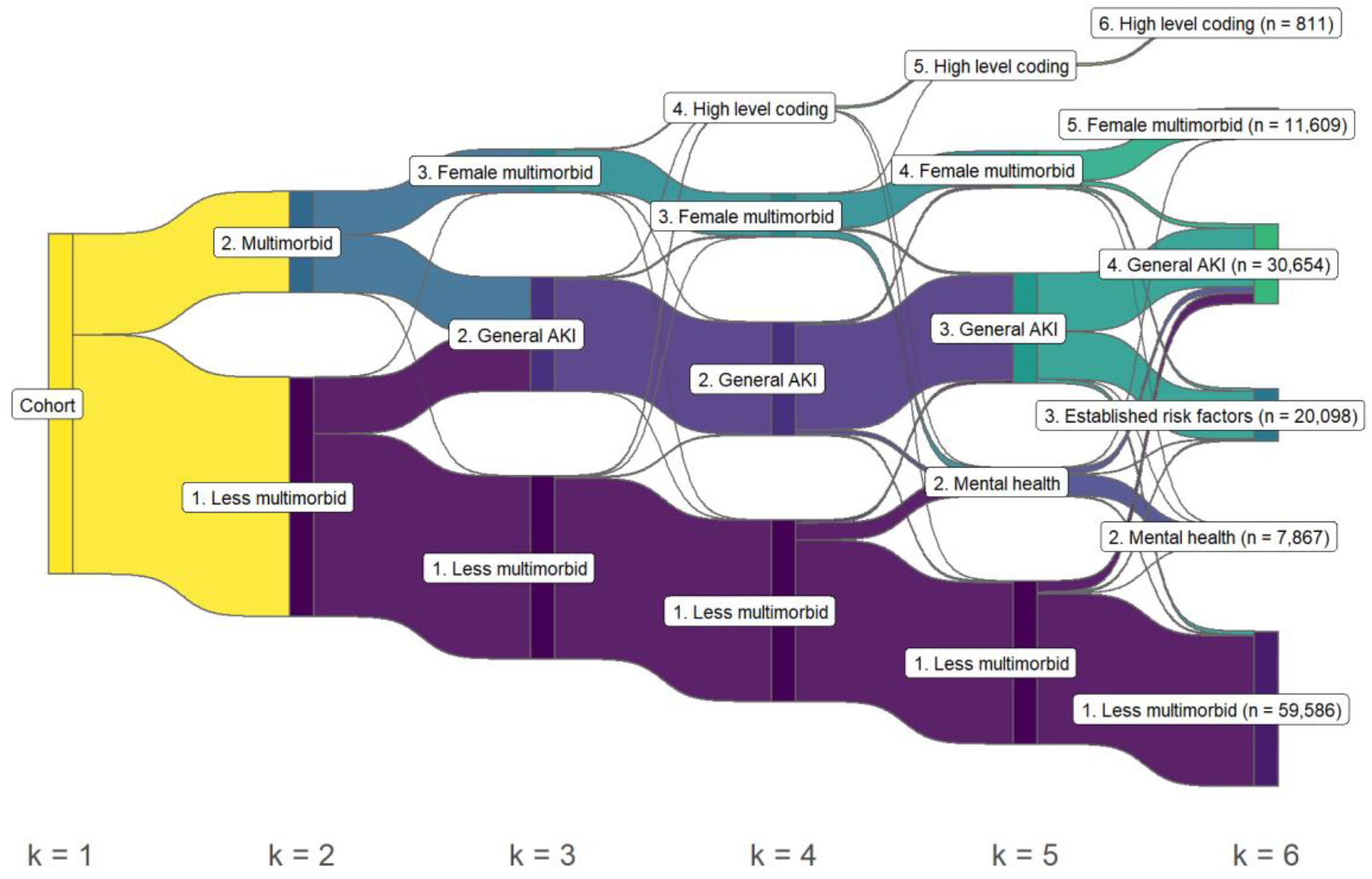
Sankey diagram of clustering assignment by k-means at each step of k for a total of 6 clusters.

We identified the following six broadly defined phenotypes from the cluster classification based on the dominant characteristics in each cluster (Figure 3, Supplementary table 5 - Supplementary table 9): Cluster 1 (Less multi-morbid phenotype): The largest cluster contained 59,586 patients defined by a younger age profile with median age of 75 (IQR: 61-85) vs. 78 (IQR: 66-86) in the cohort overall. Across the selected 17 disease codes, there were 18-50% fewer codes in this group of patients. Codes were highest for hypertensive disease (45%), rheumatism (41%), and disorders of eye and adnexa (33%).

**Figure 3:**
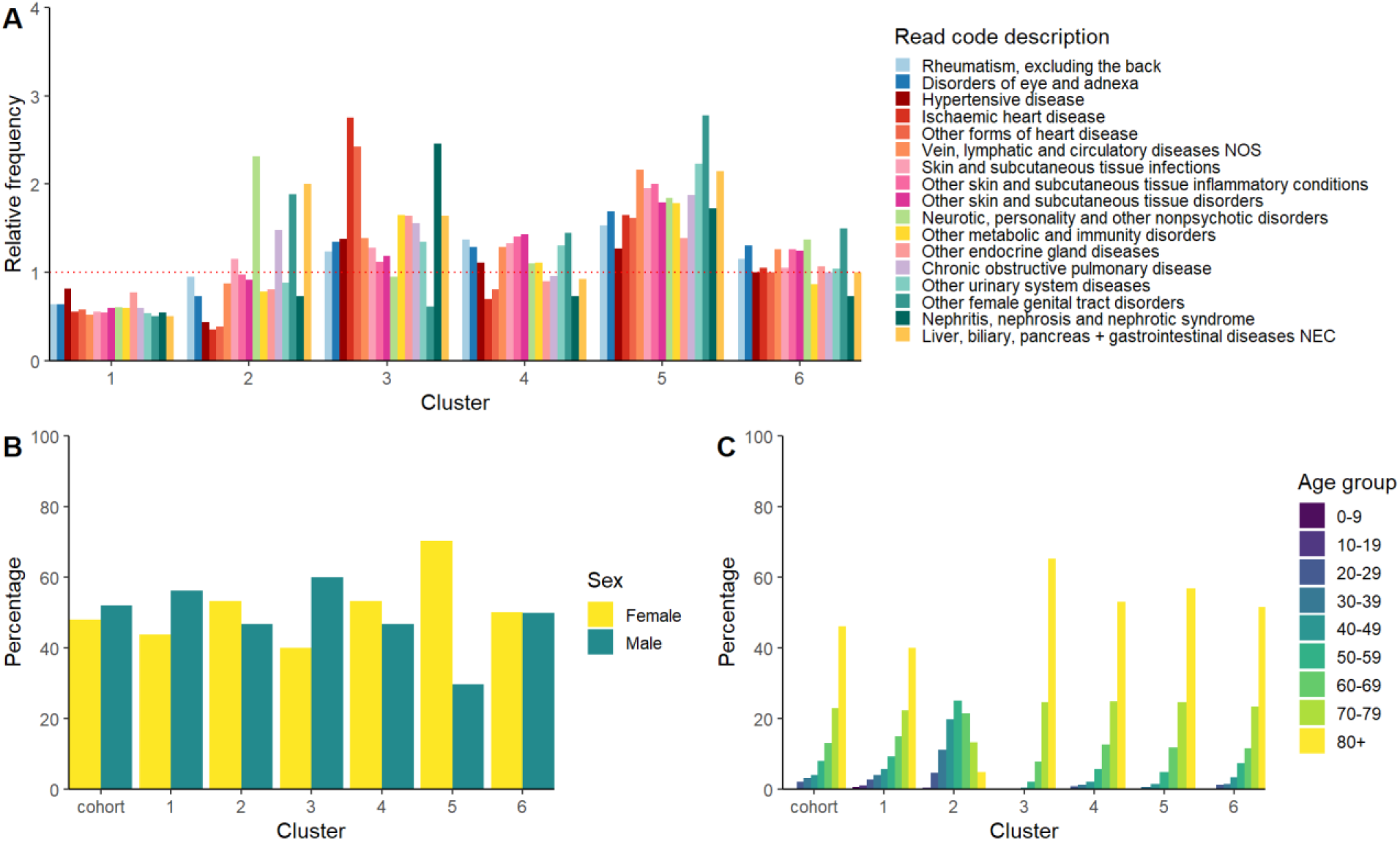
Cluster characteristics. Stratified by A) Relative frequency of cluster characteristics compared to overall cohort characteristics. B) Proportion by sex of each cluster and overall cohort. C) Percentage by age groups of each cluster and overall cohort.

Cluster 2 (Younger, mental health phenotype): The youngest cluster with median age 55 (IQR: 45-66). There were 7,867 patients in the cluster with 88% with a record of non-psychotic mental health disorders. Codes were also higher for female genital tract disorders (34%), and liver biliary, pancreas and gastrointestinal diseases (28%). 34% of patients had alcohol dependence syndrome, compared to 4% in the cohort overall (Supplementary table 10).

Cluster 3 (Established risk factors phenotype): Contained 20,098 patients, with a higher percentage of men and the oldest profile of patients with a median age of 83 (IQR: 76-88). This cluster was defined by a higher percentage of established risk factors for AKI. People had a higher proportion of cardiovascular disease codes with 2.7 times more ischemic heart disease (55%), 2.4 times more other forms of heart disease including heart failure (63%), 1.4 times more vein, lymphatic, and circulatory disease (43%), and 1.4 times more hypertensive disease (76%). Furthermore, 51% had other endocrine gland diseases including diabetes and 27% had codes for nephritis, nephrosis, and nephrotic syndrome (including acute and chronic renal failure codes), which was the highest percentage between the different clusters for both sets of codes.

Cluster 4 (General AKI phenotype): Contained 30,654 patients defined by a more typical phenotype of patient characteristics given no particular codes and conditions stood out. Few characteristics differed substantially from the overall cohort, although with a slightly higher proportion of patients with hypertensive disease (61%), rheumatism (88%), and other forms of skin and subcutaneous tissue infections, inflammatory conditions, and disorders. There were slightly more women in this group (53%) and slightly older than the cohort with a median age of 80 (IQR: 71-87).

Cluster 5 (Female multimorbid phenotype): Contained 11,609 patients with 70% female and older than the cohort overall with a median age of 81 (IQR: 73-88). Furthermore, patients in this cluster had 2.7 times more genital tract disorders (50%) such as menopausal and postmenopausal disorders, and 1.8 times more codes for non-psychotic mental disorders (70%). People in this cluster also had 2.2 times higher percentage of other urinary system diseases (58%), 2.2 times higher vein, lymphatic, and circulatory disease (67%), and 2.1 times higher percentage for liver, biliary, pancreas and gastrointestinal diseases (30%) codes than the overall cohort.

Cluster 6 (High level coding phenotype): Was a small cluster of 811 patients who were defined by a high proportion of high level diagnostic codes. For example, rather than having a code for hypertensive disease or heart failure, only a broad code is recorded such as ‘circulatory system disease’. Most frequently recorded were Read codes for digestive system disease (67%), circulatory system disease (55%), genitourinary system disease (55%), and respiratory system disease (51%) (Supplementary table 7).

#### Cluster time series

Seasonal patterns of AKI admissions were not observed for the cluster 2, 5, and 6 (Figure 4, Supplementary figure 6, Supplementary figure 7, Supplementary figure 8) while they were evident for cluster 1, 3, and 4. In addition to seasonal trends, differences in changing incidence were observed between clusters during the study period. The incidence of weekly admissions remained stable for cluster 5 and 6; increased for clusters 1, 4, and 2; declined for cluster 3 (Supplementary figure 5).

**Figure 4:**
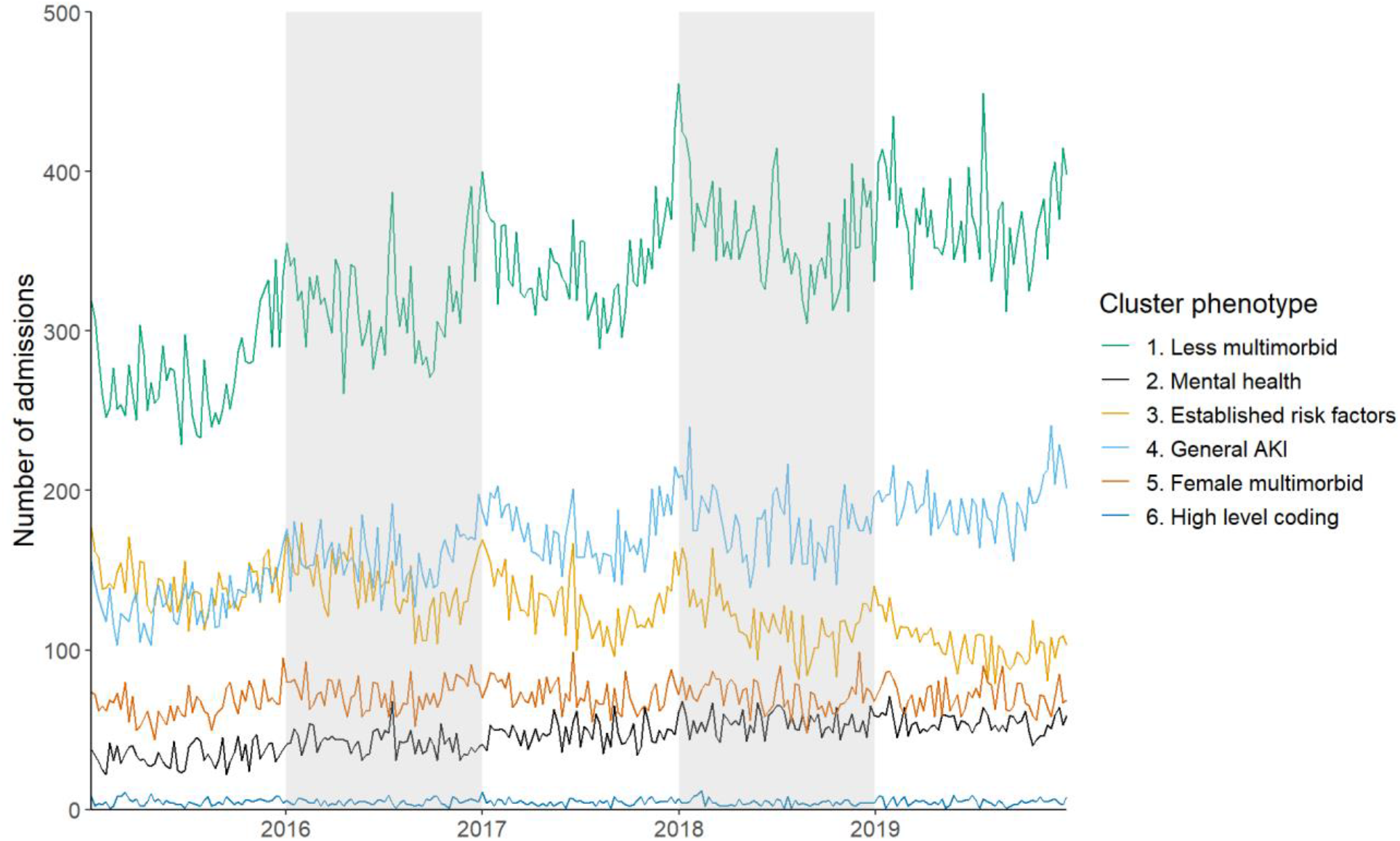
Time series of weekly AKI admissions, 2015 - 2019, England, by assigned cluster.

#### Sensitivity analysis

When conducting sensitivity analyses, the same phenotypes were identified as the primary analysis when 1) we restricted the cohort to those coded with N17 only 2) we restricted the cohort to those who had AKI recorded only in a primary diagnostic position and separately for those who had AKI recorded only in a secondary diagnostic position, 3) when we restricted the cohort to those who were diagnosed on admission (day 0), and 4) when setting random seeds for reproducibility.

We conducted a sensitivity analysis of 5) the number of dimensions included in the cluster analysis, and included 461 dimensions following MCA; equivalent to covering 70% of the variance explained in the dataset (Supplementary figure 9). Five cluster phenotypes remained the same when increasing the number of dimensions. One cluster, the mental health phenotype, was replaced with a small cluster (129 patients) of patients with musculoskeletal or connective tissue diseases.

## Discussion

Our results demonstrate that admissions involving AKI in England between 2015-2019 show a seasonal pattern with the highest peaks in December/January and further increases in June/July, coinciding with heatwaves. Admissions for people aged over 80 years showed the greatest winter seasonal increases, as well as those where AKI was diagnosed on admission suggesting the onset and cause may have been community acquired.

Using unsupervised ML clustering to generate hypotheses in a data driven approach, we identified six phenotypes of AKI admissions, of which three demonstrated marked winter seasonality. These clusters were characterised by a general AKI phenotype, those with established risk factors phenotype, and those with a younger, less multi-morbid phenotype. Using clustering methods to describe phenotypes begins to hypothesise the different profiles of patients potentially predisposed to an increased risk of AKI in the winter.

## Results in context

The observed seasonal increase of AKI in the winter months in England was consistent with previous studies which found increases in AKI reports and RRT use in the UK, and AKI admissions in Japan (4–6). However summer increases in AKI were not reported in these studies. These studies were not analysed on a weekly time scale and may not have been able to detect acute increases in admissions. Similar to findings in Japan, AKI admissions were most common in the elderly, and those diagnosed on admission, suggestive of community-acquired AKI (5). However, where pneumonia, UTIs, and sepsis were the most common primary diagnosis categories (where AKI was a secondary code) in this study, in Japan the most common admissions categories were cardiovascular and pulmonary disease. These differences may be a result of different coding practices and interpretations of primary admissions and not necessarily the underlying aetiology of AKI. The acute rise in AKI admissions we observed in the summer during heatwave alerts aligns with evidence linking increased ambient temperatures to an increased risk of AKI (8,22–28). This is an important observation given the current and increasing future impact of climate change.

Our study shows that people diagnosed with AKI have a complex multi-morbid profile and potentially have a number of mechanisms which may increase their risk of AKI, especially in winter. This is in keeping with the picture described by Philips et al, in which they found that seasonal increases in AKI affected most major medical specialities, suggesting a number of mechanisms through which AKI may increase in the winter (4).

Comparison of how the phenotypes identified in this work compare to other cluster classification studies is challenging given the large heterogeneity in approaches (29). Different methods of clustering, features included, dimension reduction techniques, and method of interpreting phenotypes (quantitative vs. qualitative) contributes to the heterogeneity in the characterisation of AKI phenotypes. Furthermore studies phenotype different subgroups of AKI such as those based on serum creatinine trajectories, severity, or biomarkers (all unavailable in this study) which further differentiate the clusters characterised from general AKI attendance (29). For example, Xu et al. used deep learning methods to characterise phenotypes of AKI patients in a critical care unit in Israel (30). The phenotypes they identified were mild, moderate, and severe kidney dysfunction which was associated with AKI stage 1, 2, and 3 respectively. Unlike our study, they found no comorbidities or demographic features defined the phenotypes identified.

## Limitations

Our study represents the most detailed examination of the seasonality of AKI in England to date. Using an unsupervised ML approach, we incorporated an unprecedented amount of data in order to be data-driven and hypothesis free and describe an objective picture of seasonal trends. However, there were several limitations to this approach. Firstly, only categorical features were included as part of phenotyping AKI (presence or absence of disease codes only). This excludes further clinical characteristics such as biomarkers, measures to determine severity of AKI, duration of AKI, or medication which could further contribute to the phenotype of AKI patients, although many of these features are not available in routine data. While the inclusion of the full clinical picture of patients with AKI at the scale needed to use machine learning may be challenging, phenotyping only diagnosis codes may bias the clinical picture and warrants caution in how these clusters are interpreted.

A further limitation was that a low proportion of the variance was explained in each dimension following dimension reduction, suggesting each variable contributes only a small amount of the variance in the data. The sensitivity analysis, which accounted for 70% of the variance, did not alter five of the cluster phenotypes, indicating that the clusters identified through the primary analysis may be stable despite being based on only a few dimensions.

While the use of Read codes enabled the examination of many diagnoses to describe clusters, it could have introduced biases in how clusters are formed due to large variation in the sensitivity and specificity of different codes. For example, using diagnostic codes alone underestimates the prevalence of CKD in CPRD (31,32) and this may have impacted on cluster formation, reducing their external validity. Assessment of the sensitivity and specificity of all 850 codes included would be challenging and presents an important limitation of an unsupervised approach to clustering. While an approach that more accurately characterises underlying comorbidities could produce clusters with higher external validity, it would likely necessitate a targeted approach that is not entirely hypothesis-free. Heterogeneity in the sensitivity and specificity of disease codes also applies for ICD-10 codes recorded in HES, with changes over time, and therefore warrants caution in interpreting longer-term trends (32,33).

### Interpretation and future studies

Our analysis of the time series data show that there are likely seasonal factors that lead to increases in AKI which is important for planning of health care services (such as surges in renal replacement therapy). AKI is a common syndrome which can lead to serious long term adverse outcomes and further evidence of seasonal increases warrants further attention of identifying which triggers, such as infectious diseases, account for the most burden. There is strong evidence of individual level associations of developing AKI following infections (7), and further studies are needed to establish whether this translates to population level drivers of AKI trends. In addition, it would be beneficial to quantify the patients at high risk, temperature triggers and burden of heat-related AKI to enable planning of appropriate responses in a changing climate.

Some individuals are more likely to be affected by winter-related triggers than others. This may be due to their underlying comorbidity profile (such as age, severity of CKD, or differences in drug therapy. However, our results highlight that some individuals who develop AKI in winter have lower levels of comorbidities. This suggests that there may be interventions that reduce the risk of seasonal AKI such as identifying those at highest risk and ensuring vaccine uptake, optimisation of medications management, or increased provision of virtual clinics to improve management of long-term conditions.

To build on our study, alternative clustering methodologies like Guassian mixture models, or supervised classification methods using known predictive AKI features could be applied. Further stratification of AKI as proposed by Vaara et al. by serum creatinine trajectories or severity of AKI may further disentangle possible aetiologies of AKI phenotypes (29). This could be achieved by the inclusion of secondary care data in defining clusters.

In conclusion, our results demonstrate how AKI incidence in England has a distinct winter and summer (heat-related) seasonal pattern which has important implications on healthcare provision planning, public health, and clinical practice.

## Supporting information

seasonality_aki_supplement_v1.0

## Data Availability

Data was provided by CPRD under terms of the LSHTM license and can be requested from CPRD. Data management and analysis code are available online at https://github.com/ehr-lshtm/acute_kidney_injury_seasonality_ML.

https://www.cprd.com/

## Contributors

HB, FS, LT, RE contributed to the conception, design, analysis and interpretation of the data for the work. HB, AS, JM contributed to the acquisition and analysis of the data including developing and reviewing code. All authors contributed to revision of the manuscript, approved the final manuscript, and agreed to be accountable for all aspects of the work and publication.

## Data sharing statement

Access to data is available on request to the Clinical Practice Research Datalink. Data management and analysis code are available online at https://github.com/ehr-lshtm/acute_kidney_injury_seasonality_ML.

## Acknowledgements

We would like to acknowledge the funding support from the National Institute for Health and Care Research (NIHR) Health Protection Research Unit (HPRU) in Modelling and Health Economics, a partnership between UK Health Security Agency (UKHSA), Imperial College London, and London School of Hygiene and Tropical Medicine. The views expressed in the study are those of the authors and not necessarily those of the National Health Service, NIHR, UK Department of Health or UKHSA. The views and opinions expressed herein are the authors’ own and do not necessarily state or reflect those of European Centre for Disease Prevention and Control (ECDC). We would also like to acknowledge Professor Elizabeth Williamson and Dr. Thomas Cowling for their invaluable advice.

