## Supplementary material for "Seasonality of acute kidney injury phenotypes in England: an unsupervised machine learning classification study of electronic health records": seasonality_aki_supplement_v1.0

Supplementary table 1: Acute kidney injury ICD10 codes

| ICD10 code | Code description |
|---|---|
| N17.0 | Acute renal failure with tubular necrosis |
| N17.1 | Acute renal failure with acute cortical necrosis |
| N17.2 | Acute renal failure with medullary necrosis |
| N17.8 | Other acute renal failure |
| N17.9 | Acute renal failure, unspecified |
| N19 | Unspecified kidney failure |

Supplementary table 2: Diagnosis Read codes included for cluster classification

| Chapter Read code | Description |
|---|---|
| A.... | Infectious and parasitic diseases |
| B.... | Neoplasms |
| C.... | Endocrine, nutritional, metabolic and immunity disorders |
| D.... | Diseases of blood and blood-forming organs |
| E.... | Mental disorders |
| F.... | Nervous system and sense organ diseases |
| G.... | Circulatory system diseases |
| H.... | Respiratory system diseases |
| J.... | Digestive system diseases |
| K.... | Genitourinary system diseases |
| L.... | Complications of pregnancy, childbirth and the puerperium |
| M.... | Skin and subcutaneous tissue diseases |
| N.... | Musculoskeletal and connective tissue diseases |
| P.... | Congenital anomalies |
| Q.... | Perinatal conditions |
| R.... | [D]Symptoms, signs and ill-defined conditions |
| S.... | Injury and poisoning |
| T.... | Causes of injury and poisoning |
| U.... | [X]External causes of morbidity and mortality |

Supplementary table 3: Diagnosis Read codes excluded for cluster classification

| Chapter Read code | Description |
|---|---|
| 0.... | Occupations |
| 1.... | History / symptoms |
| 2.... | Examination / signs |
| 3.... | Diagnostic procedures |
| 4.... | Laboratory procedures |
| 5.... | Radiology/physics in medicine |
| 6.... | Preventive procedures |
| 7.... | Operations, procedures, sites |
| 8.... | Other therapeutic procedures |
| 9.... | Administration |
| Z.... | Unspecified conditions |
| a.... | Gastro-intestinal drugs |
| b.... | Cardiovascular drugs |
| c.... | Respiratory drugs |
| d.... | Central nervous system drugs |
| e.... | Drugs used in infections |
| f.... | Endocrine drugs |
| g.... | Obs/gynae/uti drugs |
| h.... | Chemotherapy/immunosuppressant drugs |
| i.... | Haematology/dietetic drugs |
| j.... | Musculoskeletal drugs |
| k.... | Eye drugs |
| m.... | Skin drugs |
| n.... | Immunology drugs and vaccines |
| o.... | Anaesthetic drugs |
| p.... | Appliances & reagents etc |
| q.... | Incontinence appliances |
| r.... | Appliances & reagents etc(2) |
| s.... | Stoma appliances |
| u.... | Contrast media |
| y.... | Drug release administration |

Supplementary table 4: Top 20 primary diagnostic ICD-10 codes where AKI was a secondary code during the admission

| ICD10 chapter diagnosis | ICD10 chapter code | n | percentage |
|---|---|---|---|
| Bronchopneumonia, unspecified | J18 | 18,429 | 11% |
| Other sepsis | A41 | 14,335 | 8% |
| Urinary tract infection, site not specified | N39 | 12,128 | 7% |
| Heart failure | I50 | 6,933 | 4% |
| Other chronic obstructive pulmonary disease | J44 | 3,757 | 2% |
| Other gastroenteritis and colitis of infectious and unspecified origin | A09 | 3,705 | 2% |
| Fracture of femur | S72 | 3,419 | 2% |
| Cellulitis | L03 | 3,304 | 2% |
| Unspecified acute lower respiratory infection | J22 | 3,207 | 2% |
| Other symptoms and signs involving the nervous and musculoskeletal systems | R29 | 2,918 | 2% |
| Acute myocardial infarction | I21 | 2,887 | 2% |
| Other disorders of fluid, electrolyte and acid-base balance | E87 | 2,350 | 1% |
| Pneumonitis due to solids and liquids | J69 | 2,120 | 1% |
| Obstructive and reflux uropathy | N13 | 2,062 | 1% |
| Non-insulin-dependent diabetes mellitus | E11 | 1,808 | 1% |
| Cerebral infarction | I63 | 1,761 | 1% |
| Haematemesis | K92 | 1,719 | 1% |
| Type 1 diabetes mellitus | E10 | 1,693 | 1% |
| Atrial fibrillation and flutter | I48 | 1,588 | 1% |
| Alcoholic liver disease | K70 | 1,511 | 1% |

Supplementary figure 1. Normalised AKI admissions in HES-linked CPRD 2015-2029. Time series of weekly AKI admissions, 2015 - 2019, England total and by A) sex of patients B) age group C) day AKI code was recorded during the admission D) diagnostic position of AKI record E) primary diagnosis where AKI code was recorded during the admission (primary diagnoses displayed make up 36% of all primary diagnoses recorded).

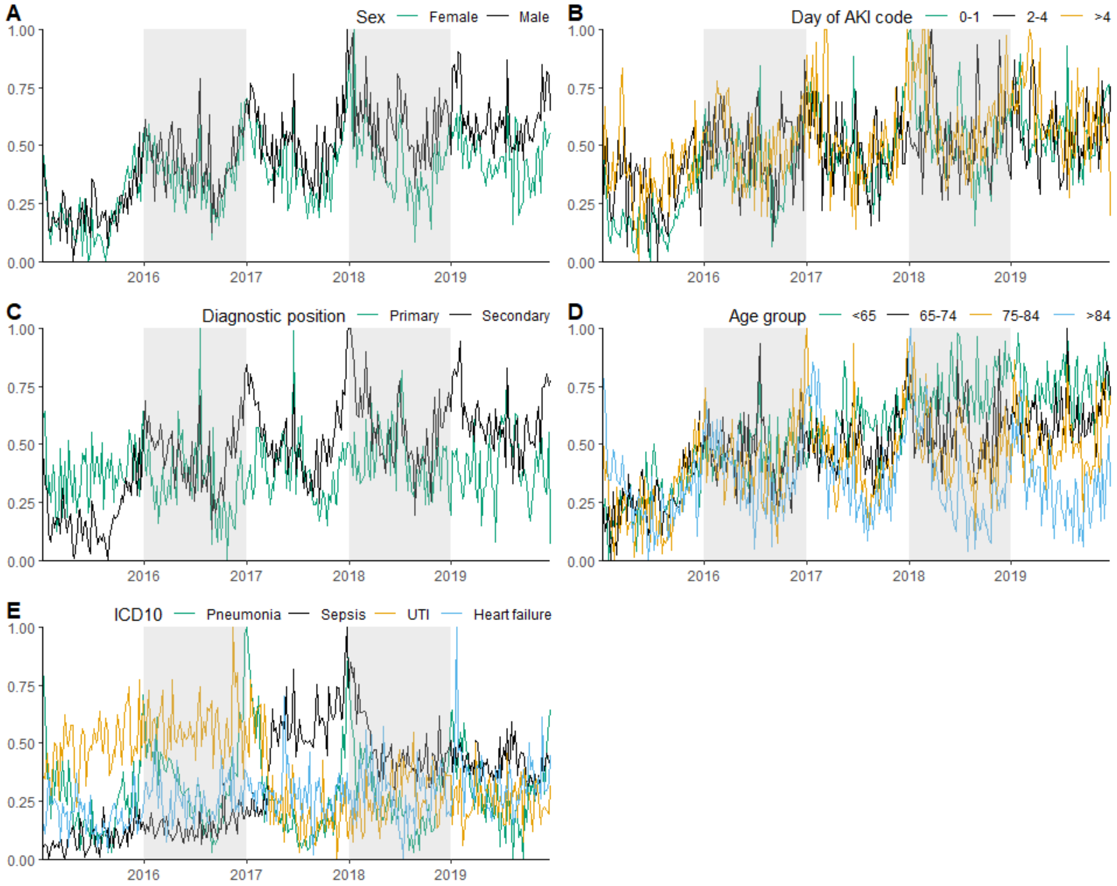

Supplementary figure 2. Time series of weekly AKI admissions, 2015 - 2019, England by A) number of admissions where AKI was recorded in a primary diagnostic position B) number of admissions where AKI was recorded in a secondary diagnostic position C) diagnostic position of AKI record and number of admissions on a normalised scale. Red shaded areas denote the periods in which heat waves were declared by the Met Office in the UK.

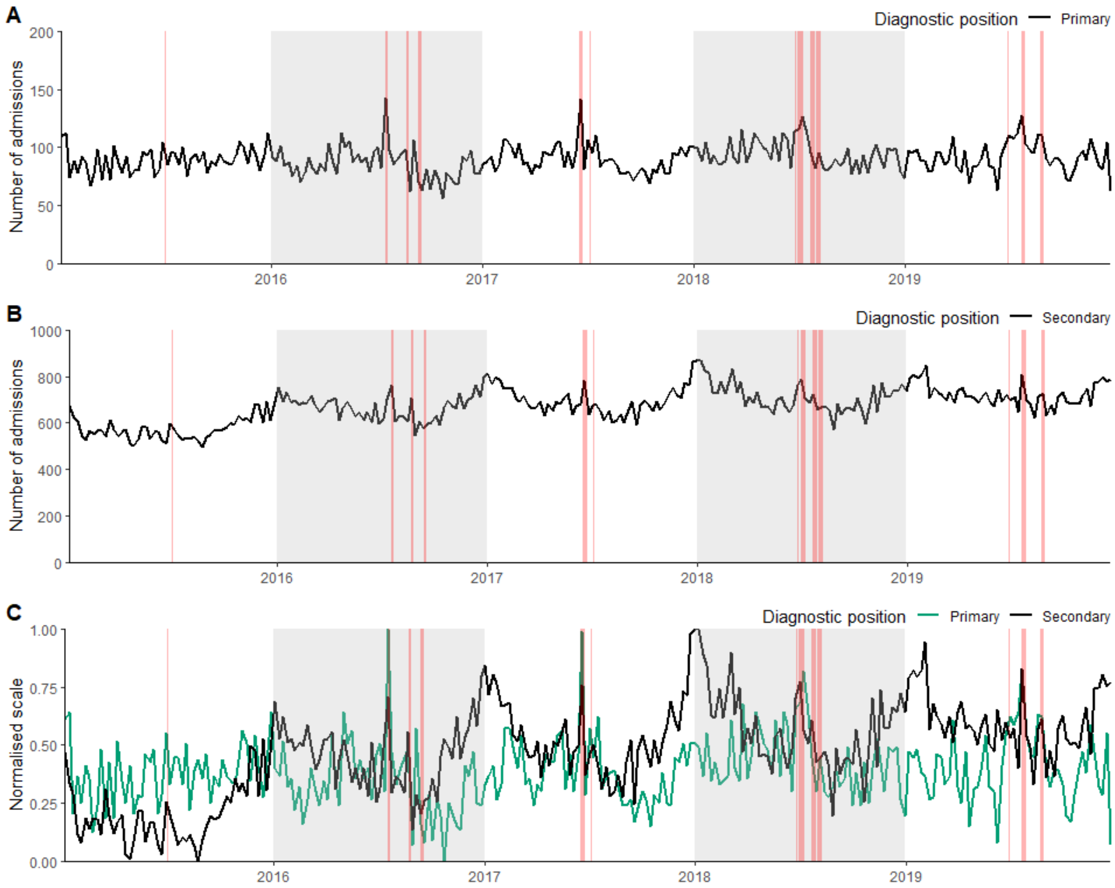

Supplementary figure 3. Scree plot of number of dimensions and percentage of explained variance.

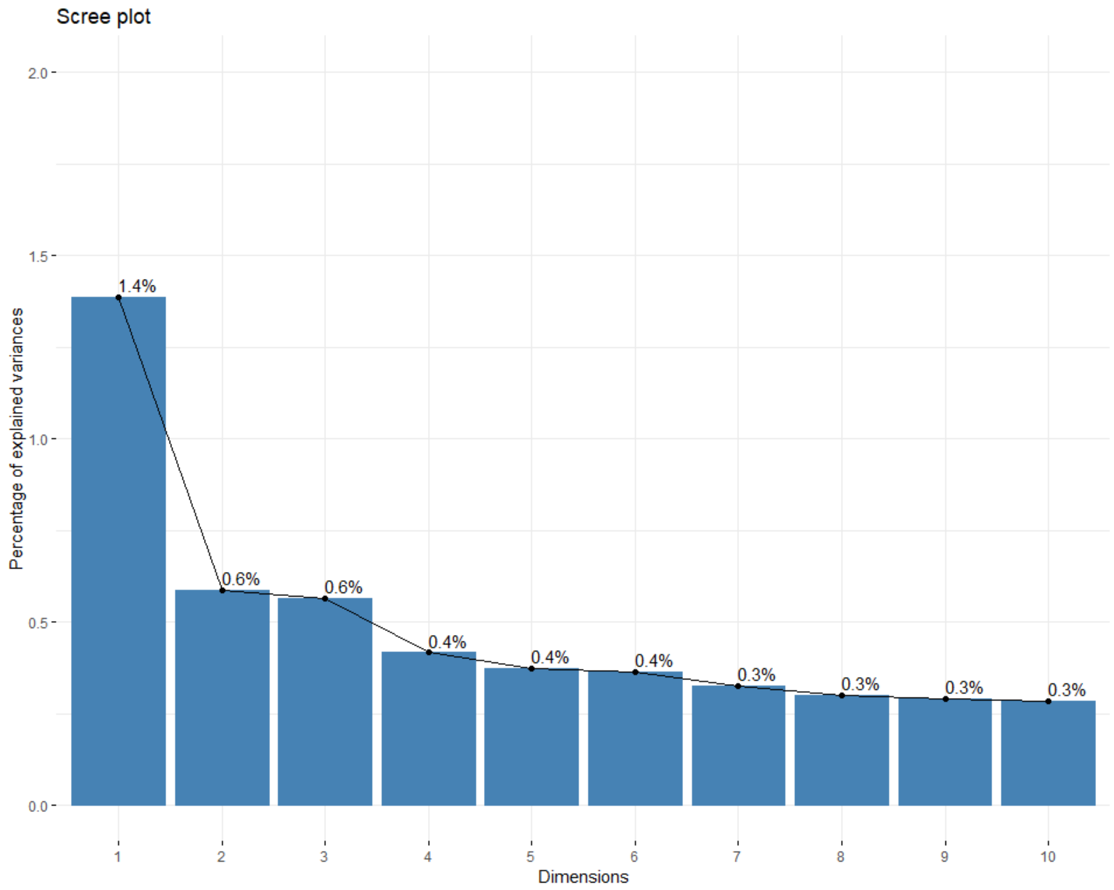

Supplementary figure 4. NBClust package output of five runs. All indices were run except GAP, Gamma, Gplus and Tau.

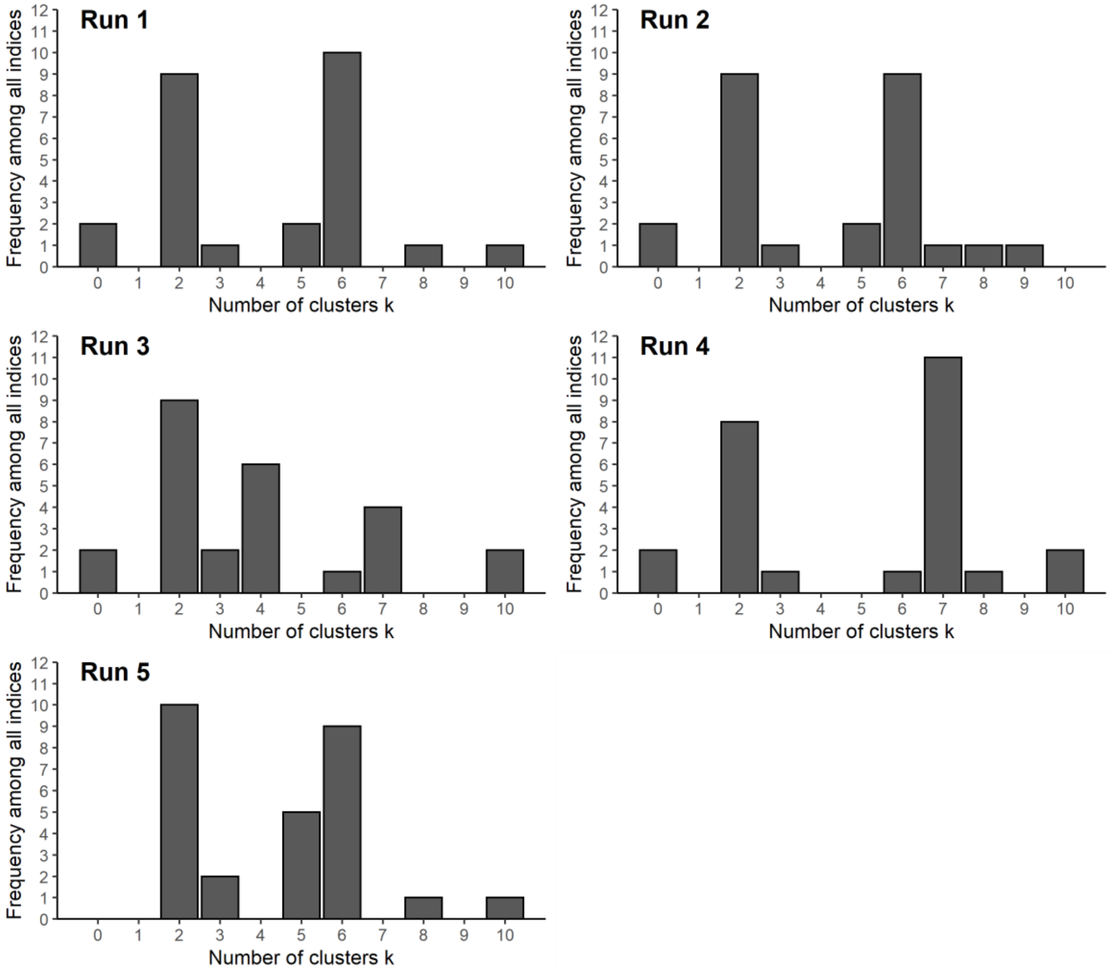

Supplementary figure 5. 2D visualization of cluster assignment k = 6

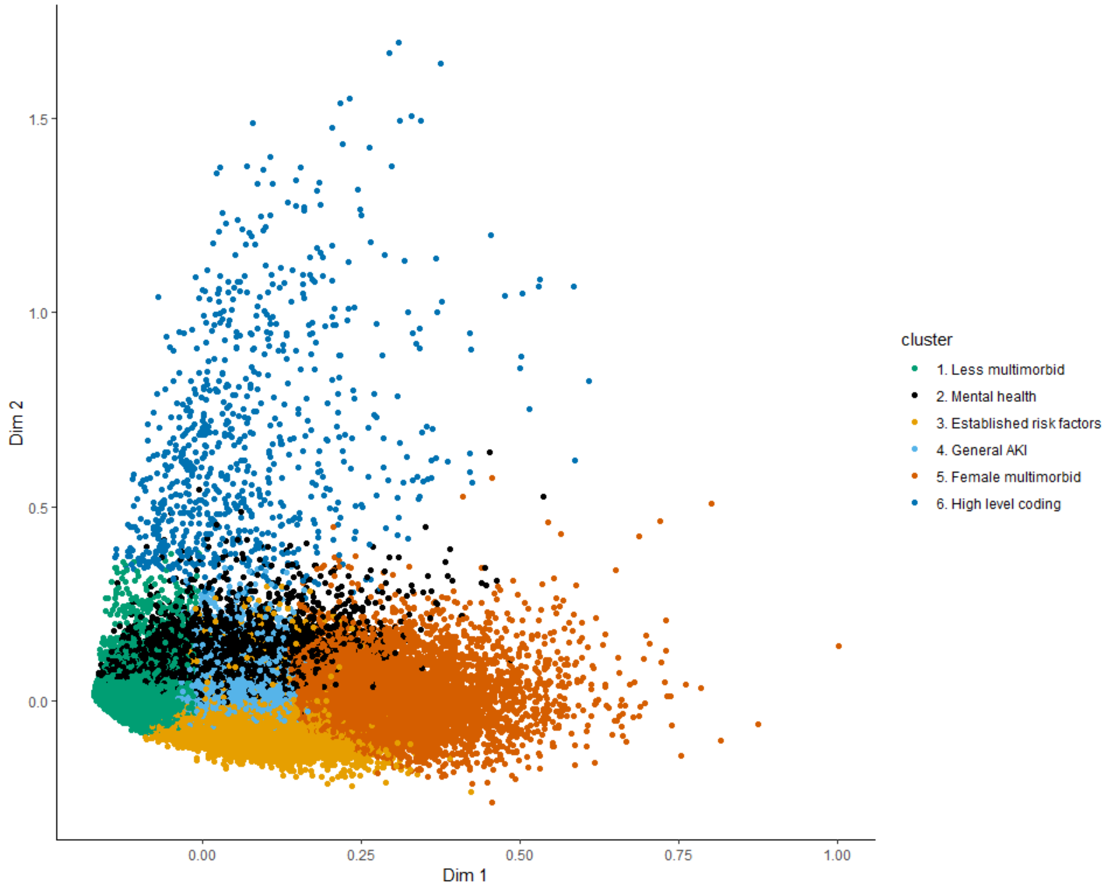

Supplementary figure 6. Normalised time series of weekly AKI admissions, 2015 - 2019, England, by assigned cluster

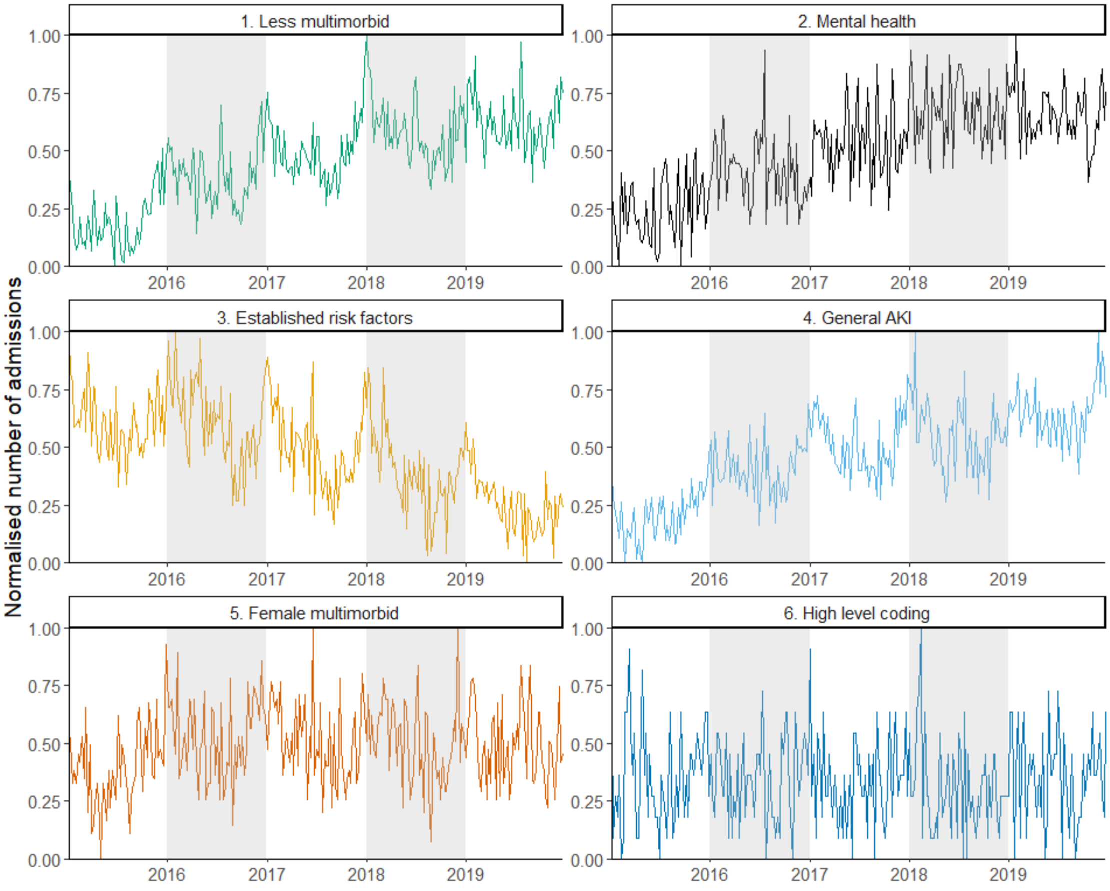

Supplementary figure 7. Time series of weekly AKI admissions, 2015 - 2019, England, by assigned cluster and epidemiological week

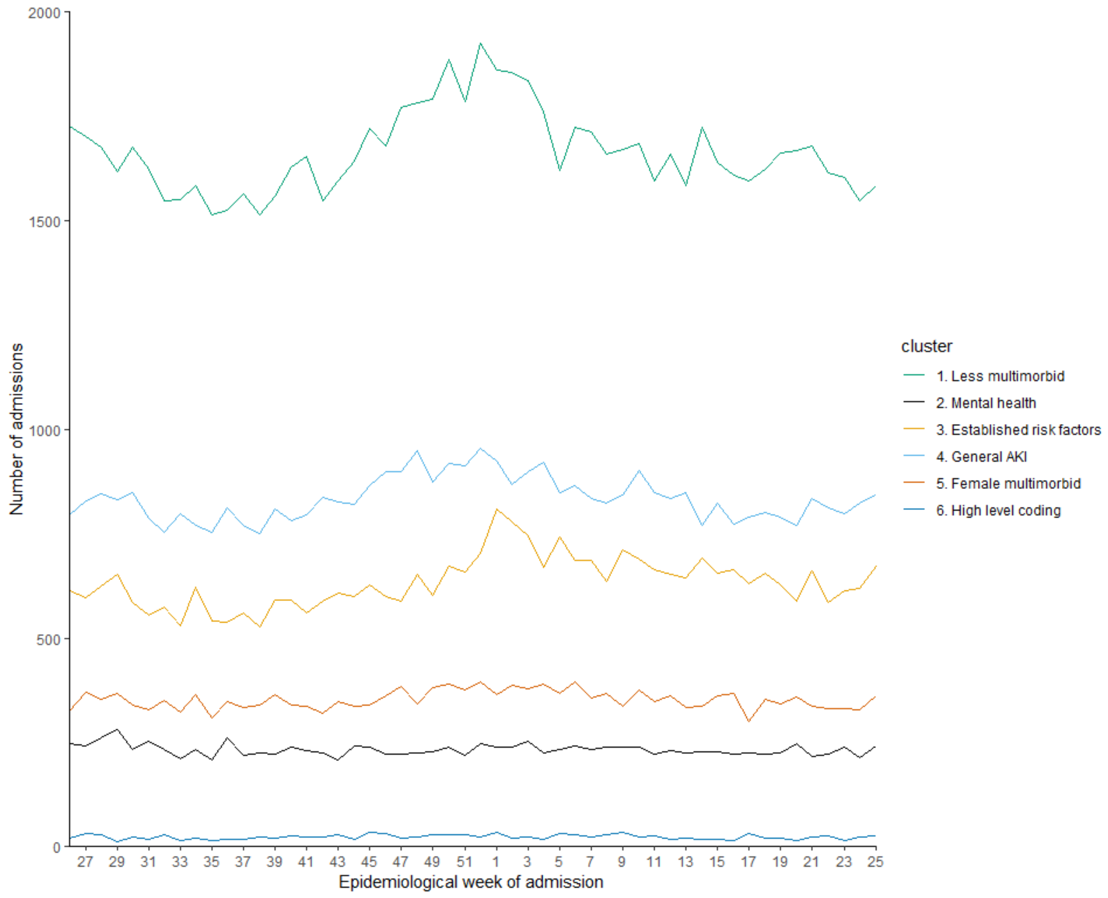

Supplementary figure 8. Time series of weekly AKI admissions, 2015 - 2019, England, by year and assigned cluster, epidemiological week

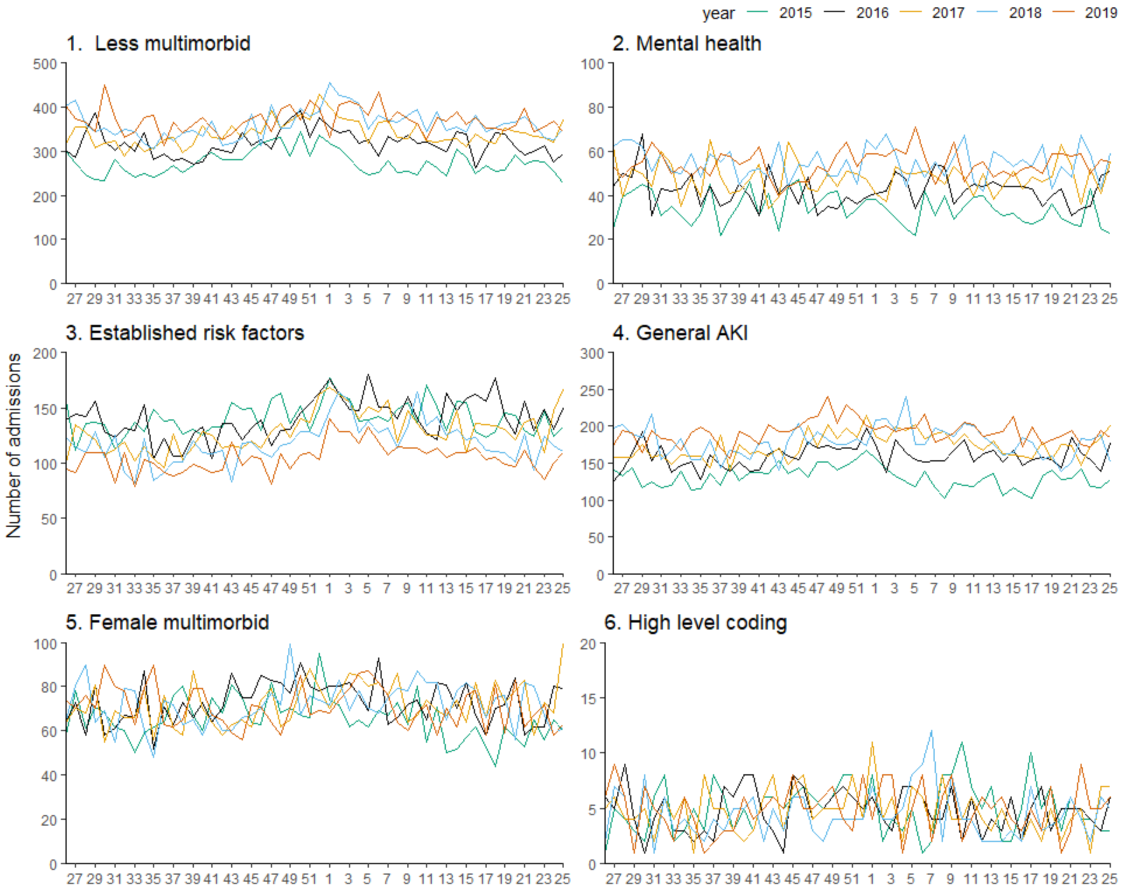

Supplementary figure 9. Sankey diagram of clustering assignment by k-means for 6 clusters for 5 dimensions and 461 dimensions to show the movement of assignments. As the dimensions increased, the mental health cluster was re-assigned to other groups and replaced with a small cluster of patients with musculoskeletal or connective tissue diseases.

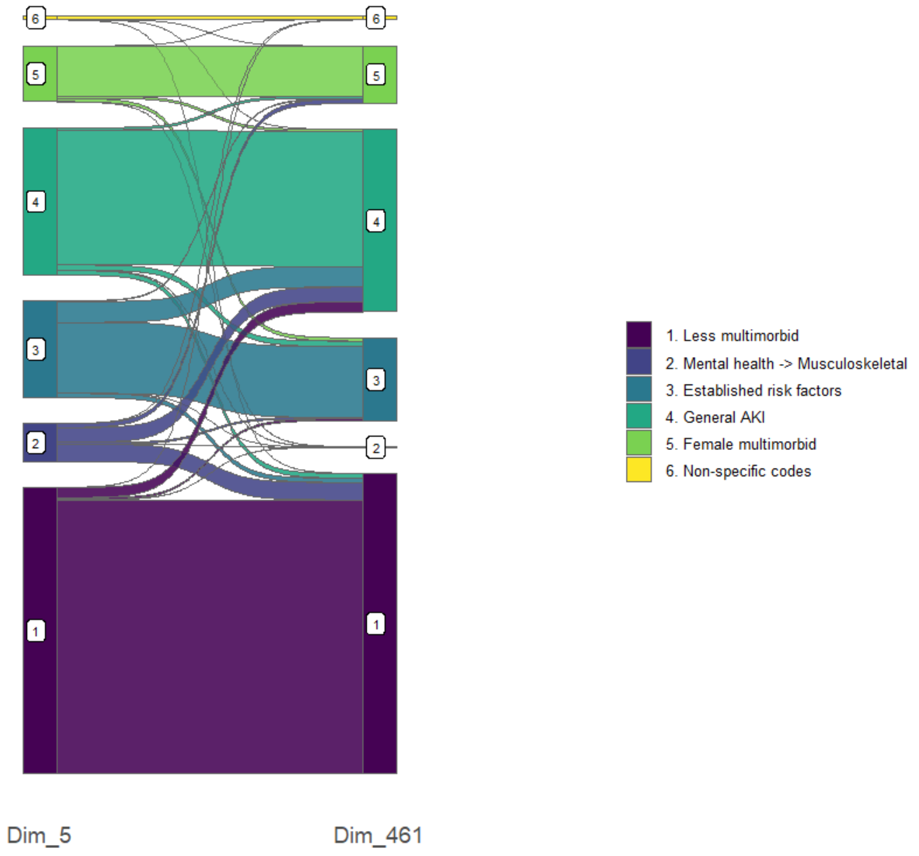

Supplementary table 5. Summary table of cluster characteristics (Percentage with the characteristic in the cohort/cluster unless otherwise specified)

| Variable | cohort | 1 | 2 | 3 | 4 | 5 | 6 |
|---|---|---|---|---|---|---|---|
| Sex | | | | | | | |
| Female | 48% | 44% | 53% | 40% | 53% | 70% | 50% |
| Male | 52% | 56% | 47% | 60% | 47% | 30% | 50% |
| Age | | | | | | | |
| Median (IQR) | 78 (66-86) | 75 (61-85) | 55 (45-66) | 83 (76-88) | 80 (71-87) | 81 (73-88) | 80 (69.5-87) |
| Chapter headings | | | | | | | |
| N2 (Rheumatism, excluding the back) | 64% | 41% | 61% | 79% | 88% | 98% | 74% |
| F4 (Disorders of eye and adnexa) | 52% | 33% | 38% | 70% | 67% | 88% | 68% |
| G2 (Hypertensive disease) | 55% | 45% | 24% | 76% | 61% | 70% | 55% |
| G3 (Ischaemic heart disease) | 20% | 11% | 7% | 55% | 14% | 33% | 21% |
| G5 (Other forms of heart disease) | 26% | 15% | 10% | 63% | 21% | 42% | 26% |
| G8 (Vein, lymphatic and circulatory diseases NOS) | 31% | 16% | 27% | 43% | 40% | 67% | 39% |
| M0 (Skin and subcutaneous tissue infections) | 40% | 22% | 46% | 51% | 53% | 78% | 42% |
| M1 (Other skin and subcutaneous tissue inflammatory conditions) | 42% | 23% | 41% | 47% | 59% | 84% | 53% |
| M2 (Other skin and subcutaneous tissue disorders) | 49% | 29% | 45% | 58% | 70% | 88% | 61% |
| E2 (Neurotic, personality and other nonpsychotic disorders) | 38% | 23% | 88% | 36% | 42% | 70% | 52% |
| C3 (Other metabolic and immunity disorders) | 37% | 22% | 29% | 61% | 41% | 66% | 32% |
| C1 (Other endocrine gland diseases) | 31% | 24% | 25% | 51% | 28% | 43% | 33% |
| H3 (Chronic obstructive pulmonary disease) | 25% | 15% | 37% | 39% | 24% | 47% | 25% |
| K1 (Other urinary system diseases) | 26% | 14% | 23% | 35% | 34% | 58% | 27% |
| K5 (Other female genital tract disorders) | 18% | 9% | 34% | 11% | 26% | 50% | 27% |
| K0 (Nephritis, nephrosis and nephrotic syndrome) | 11% | 6% | 8% | 27% | 8% | 19% | 8% |
| J6 (Liver, biliary, pancreas + gastrointestinal diseases NEC) | 14% | 7% | 28% | 23% | 13% | 30% | 14% |

Supplementary table 5. Summary table of cluster characteristics (Percentage with the characteristic in the cohort/cluster unless otherwise specified). * denotes suppression due to small numbers.

| Age group | cohort | 1 | 2 | 3 | 4 | 5 | 6 |
|---|---|---|---|---|---|---|---|
| 0-9 | 0% (290) | 0% (288) | 0% (0) | * | * | * | * |
| 10-19 | 0% (618) | 1% (549) | 0% (26) | * | * | * | * |
| 20-29 | 2% (2255) | 3% (1602) | 4% (354) | * | 1% (260) | * | * |
| 30-39 | 3% (3634) | 4% (2308) | 11% (875) | * | 1% (358) | 1% (59) | * |
| 40-49 | 4% (5745) | 6% (3336) | 20% (1550) | 0% (69) | 2% (600) | 1% (164) | 3% (26) |
| 50-59 | 8% (10204) | 9% (5508) | 25% (1972) | 2% (391) | 6% (1708) | 5% (566) | 7% (59) |
| 60-69 | 13% (17392) | 15% (8879) | 21% (1683) | 8% (1552) | 12% (3829) | 12% (1355) | 12% (94) |
| 70-79 | 23% (29915) | 22% (13296) | 13% (1031) | 25% (4943) | 25% (7610) | 25% (2846) | 23% (189) |
| 80+ | 46% (60572) | 40% (23820) | 5% (376) | 65% (13113) | 53% (16248) | 57% (6596) | 52% (419) |
| Median (IQR) | 78 (66-86) | 75 (61-85) | 55 (45-66) | 83 (76-88) | 80 (71-87) | 81 (73-88) | 80 (69.5-87) |

Supplementary table 7: Top 15 codes of cohort and clusters at Read code chapter level 2 (Percentage with the code in the cohort/cluster).

| | cohort | 1 | 2 | 3 | 4 | 5 | 6 |
|---|---|---|---|---|---|---|---|
| 1 | Rheumatism, excluding the back (64%) | Hypertensive disease (45%) | Neurotic, personality and other nonpsychotic disorders (88%) | [D]Symptoms (80%) | Rheumatism, excluding the back (88%) | [D]Symptoms (99%) | Rheumatism, excluding the back (74%) |
| 2 | [D]Symptoms (64%) | Rheumatism, excluding the back (41%) | [D]Symptoms (73%) | Rheumatism, excluding the back (79%) | [D]Symptoms (85%) | Rheumatism, excluding the back (98%) | [D]Symptoms (74%) |
| 3 | Hypertensive disease (55%) | [D]Symptoms (39%) | [X]Mental and behavioural disorders (64%) | Hypertensive disease (76%) | Arthropathies and related disorders (72%) | Arthropathies and related disorders (93%) | Disorders of eye and adnexa (68%) |
| 4 | Disorders of eye and adnexa (52%) | Disorders of eye and adnexa (33%) | Rheumatism, excluding the back (61%) | Disorders of eye and adnexa (70%) | Other skin and subcutaneous tissue disorders (70%) | Vertebral column syndromes (91%) | Skin and subcutaneous tissue diseases (68%) |
| 5 | Arthropathies and related disorders (52%) | Arthropathies and related disorders (30%) | Vertebral column syndromes (47%) | Arthropathies and related disorders (66%) | Vertebral column syndromes (69%) | Disorders of eye and adnexa (88%) | Digestive system diseases (67%) |
| 6 | Other skin and subcutaneous tissue disorders (49%) | Other skin and subcutaneous tissue disorders (29%) | Skin and subcutaneous tissue infections (46%) | Other forms of heart disease (63%) | Disorders of eye and adnexa (67%) | Other skin and subcutaneous tissue disorders (88%) | Arthropathies and related disorders (63%) |
| 7 | Vertebral column syndromes (48%) | Diseases of the ear and mastoid process (26%) | Other skin and subcutaneous tissue disorders (45%) | Other metabolic and immunity disorders (61%) | Diseases of the ear and mastoid process (65%) | Other skin and subcutaneous tissue inflammatory conditions (84%) | Other skin and subcutaneous tissue disorders (61%) |
| 8 | Diseases of the ear and mastoid process (45%) | Vertebral column syndromes (25%) | Other skin and subcutaneous tissue inflammatory conditions (41%) | Other skin and subcutaneous tissue disorders (58%) | Hypertensive disease (61%) | Diseases of the ear and mastoid process (83%) | Diseases of the ear and mastoid process (56%) |
| 9 | Other skin and subcutaneous tissue inflammatory conditions (42%) | Other endocrine gland diseases (24%) | Diseases of the ear and mastoid process (40%) | Vertebral column syndromes (56%) | Other skin and subcutaneous tissue inflammatory conditions (59%) | Skin and subcutaneous tissue infections (78%) | Vertebral column syndromes (56%) |
| 10 | Skin and subcutaneous tissue infections (40%) | Neurotic, personality and other nonpsychotic disorders (23%) | Oesophageal, stomach and duodenal diseases (39%) | Ischaemic heart disease (55%) | Skin and subcutaneous tissue infections (53%) | Oesophageal, stomach and duodenal diseases (71%) | Circulatory system diseases (55%) |
| 11 | Neurotic, personality and other nonpsychotic disorders (38%) | Other skin and subcutaneous tissue inflammatory conditions (23%) | Arthropathies and related disorders (39%) | Other endocrine gland diseases (51%) | Neurotic, personality and other nonpsychotic disorders (42%) | Neurotic, personality and other nonpsychotic disorders (70%) | Hypertensive disease (55%) |
| 12 | Other metabolic and immunity disorders (37%) | Other metabolic and immunity disorders (22%) | Poisoning (39%) | Skin and subcutaneous tissue infections (51%) | Other metabolic and immunity disorders (41%) | Hypertensive disease (70%) | Genitourinary system diseases (55%) |
| 13 | Oesophageal, stomach and duodenal diseases (33%) | Skin and subcutaneous tissue infections (22%) | Disorders of eye and adnexa (38%) | Diseases of the ear and mastoid process (49%) | Vein, lymphatic and circulatory diseases NOS (40%) | Other diseases of the intestines and peritoneum (68%) | Other skin and subcutaneous tissue inflammatory conditions (53%) |

| | cohort | 1 | 2 | 3 | 4 | 5 | 6 |
|---|---|---|---|---|---|---|---|
| 14 | Other endocrine gland diseases (31%) | [X]Mental and behavioural disorders (16%) | Chronic obstructive pulmonary disease (37%) | Other skin and subcutaneous tissue inflammatory conditions (47%) | Oesophageal, stomach and duodenal diseases (40%) | Vein, lymphatic and circulatory diseases NOS (67%) | Neurotic, personality and other nonpsychotic disorders (52%) |
| 15 | Vein, lymphatic and circulatory diseases NOS (31%) | Vein, lymphatic and circulatory diseases NOS (16%) | Other female genital tract disorders (34%) | Oesophageal, stomach and duodenal diseases (46%) | Other diseases of the intestines and peritoneum (39%) | Other metabolic and immunity disorders (66%) | Respiratory system diseases (51%) |

Supplementary table 8: Top 15 codes of cohort and clusters at Read code chapter level 3 (Percentage with the code in the cohort/cluster).

| | cohort | 1 | 2 | 3 | 4 | 5 | 6 |
|---|---|---|---|---|---|---|---|
| 1 | Other soft tissue disorders (54%) | Essential hypertension (36%) | Neurotic disorders (51%) | Other soft tissue disorders (69%) | Other soft tissue disorders (77%) | Other soft tissue disorders (96%) | Skin and subcutaneous tissue diseases (68%) |
| 2 | Essential hypertension (45%) | Other soft tissue disorders (30%) | Other soft tissue disorders (51%) | Essential hypertension (63%) | Other and unspecified joint disorders (55%) | Other and unspecified joint disorders (82%) | Digestive system diseases (67%) |
| 3 | Other and unspecified joint disorders (36%) | Diabetes mellitus (21%) | [X]Mood - affective disorders (44%) | Diabetes mellitus (44%) | Essential hypertension (50%) | [D]General symptoms (80%) | Other soft tissue disorders (57%) |
| 4 | Other and unspecified back disorders (35%) | Other and unspecified back disorders (17%) | [D]General symptoms (41%) | Osteoarthritis and allied disorders (44%) | Other and unspecified back disorders (50%) | Other and unspecified back disorders (78%) | Circulatory system diseases (55%) |
| 5 | [D]General symptoms (33%) | Hypertensive disease (16%) | Poisoning (39%) | Other and unspecified joint disorders (42%) | [D]General symptoms (46%) | [D]Symptoms affecting skin and other integumentary tissue (68%) | Genitourinary system diseases (55%) |
| 6 | Osteoarthritis and allied disorders (31%) | Osteoarthritis and allied disorders (16%) | Other and unspecified back disorders (36%) | Cardiac dysrhythmias (41%) | Osteoarthritis and allied disorders (43%) | Osteoarthritis and allied disorders (65%) | Respiratory system diseases (51%) |
| 7 | Peripheral enthesopathies and allied syndromes (28%) | Other and unspecified joint disorders (16%) | Alcohol dependence syndrome (34%) | [D]General symptoms (40%) | Peripheral enthesopathies and allied syndromes (42%) | Peripheral enthesopathies and allied syndromes (63%) | Musculoskeletal and connective tissue diseases (51%) |
| 8 | [D]Symptoms affecting skin and other integumentary tissue (27%) | [D]General symptoms (14%) | Other and unspecified joint disorders (30%) | Other and unspecified back disorders (39%) | [D]Symptoms affecting skin and other integumentary tissue (39%) | Essential hypertension (60%) | Other and unspecified joint disorders (47%) |
| 9 | Diabetes mellitus (26%) | Peripheral enthesopathies and allied syndromes (13%) | Depressive disorder NEC (28%) | Ischaemic heart disease (34%) | Disorders of external ear (37%) | [D]Respiratory system and chest symptoms (58%) | Essential hypertension (44%) |
| 10 | Disorders of external ear (23%) | Disorders of lipoid metabolism (11%) | Asthma (26%) | Hypertensive disease (33%) | Disorders of conjunctiva (32%) | Other cervical disorders (57%) | Other and unspecified back disorders (42%) |
| 11 | Hypertensive disease (23%) | Disorders of external ear (11%) | [D]Other abdominal and pelvic symptoms (26%) | Peripheral enthesopathies and allied syndromes (33%) | Other cervical disorders (31%) | Disorders of conjunctiva (55%) | Peripheral enthesopathies and allied syndromes (41%) |
| 12 | Disorders of conjunctiva (22%) | [D]Symptoms affecting skin and other integumentary tissue (11%) | [D]Symptoms affecting skin and other integumentary tissue (24%) | Heart failure (32%) | Hypertensive disease (28%) | [D]Other abdominal and pelvic symptoms (54%) | Disorders of eye and adnexa (38%) |
| 13 | Other cellulitis and abscess (21%) | Neurotic disorders (10%) | Other specified injury (23%) | [D]Symptoms affecting skin and other integumentary tissue (32%) | Other dermatoses (28%) | Disorders of external ear (53%) | Osteoarthritis and allied disorders (37%) |

| | cohort | 1 | 2 | 3 | 4 | 5 | 6 |
|---|---|---|---|---|---|---|---|
| 14 | Other cervical disorders (20%) | Disorders of conjunctiva (10%) | Menstruation disorders (22%) | Disorders of lipoid metabolism (31%) | Contact dermatitis and other eczemas (27%) | Accidental falls (49%) | [D]General symptoms (35%) |
| 15 | [D]Respiratory system and chest symptoms (20%) | Cardiac dysrhythmias (10%) | Other cellulitis and abscess (22%) | Other cellulitis and abscess (30%) | [D]Other abdominal and pelvic symptoms (25%) | Contact dermatitis and other eczemas (47%) | Mental disorders (34%) |

Supplementary table 9: Code characteristics of cohort and clusters at Read code chapter level 2 (Percentage with the code in the cohort/cluster). Presenting 100 of 164 codes total.

| chapter | cohort | 1 | 2 | 3 | 4 | 5 | 6 |
|---|---|---|---|---|---|---|---|
| N2 (Rheumatism, excluding the back) | 64% | 41% | 61% | 79% | 88% | 98% | 74% |
| R0 ([D]Symptoms) | 64% | 39% | 73% | 80% | 85% | 99% | 74% |
| G2 (Hypertensive disease) | 55% | 45% | 24% | 76% | 61% | 70% | 55% |
| F4 (Disorders of eye and adnexa) | 52% | 33% | 38% | 70% | 67% | 88% | 68% |
| N0 (Arthropathies and related disorders) | 52% | 30% | 39% | 66% | 72% | 93% | 63% |
| M2 (Other skin and subcutaneous tissue disorders) | 49% | 29% | 45% | 58% | 70% | 88% | 61% |
| N1 (Vertebral column syndromes) | 48% | 25% | 47% | 56% | 69% | 91% | 56% |
| F5 (Diseases of the ear and mastoid process) | 45% | 26% | 40% | 49% | 65% | 83% | 56% |
| M1 (Other skin and subcutaneous tissue inflammatory conditions) | 42% | 23% | 41% | 47% | 59% | 84% | 53% |
| M0 (Skin and subcutaneous tissue infections) | 40% | 22% | 46% | 51% | 53% | 78% | 42% |
| E2 (Neurotic, personality and other nonpsychotic disorders) | 38% | 23% | 88% | 36% | 42% | 70% | 52% |
| C3 (Other metabolic and immunity disorders) | 37% | 22% | 29% | 61% | 41% | 66% | 32% |
| J1 (Oesophageal, stomach and duodenal diseases) | 33% | 16% | 39% | 46% | 40% | 71% | 40% |
| C1 (Other endocrine gland diseases) | 31% | 24% | 25% | 51% | 28% | 43% | 33% |
| G8 (Vein, lymphatic and circulatory diseases NOS) | 31% | 16% | 27% | 43% | 40% | 67% | 39% |
| J5 (Other diseases of the intestines and peritoneum) | 31% | 15% | 32% | 43% | 39% | 68% | 39% |
| G5 (Other forms of heart disease) | 26% | 15% | 10% | 63% | 21% | 42% | 26% |
| K1 (Other urinary system diseases) | 26% | 14% | 23% | 35% | 34% | 58% | 27% |
| H3 (Chronic obstructive pulmonary disease) | 25% | 15% | 37% | 39% | 24% | 47% | 25% |
| Eu ([X]Mental and behavioural disorders) | 22% | 16% | 64% | 25% | 19% | 33% | 18% |
| TC (Accidental falls) | 21% | 9% | 18% | 30% | 25% | 50% | 18% |
| G3 (Ischaemic heart disease) | 20% | 11% | 7% | 55% | 14% | 33% | 21% |
| R1 ([D]Nonspecific abnormal findings) | 20% | 11% | 14% | 33% | 22% | 41% | 26% |
| J3 (Hernia of abdominal cavity) | 18% | 9% | 14% | 29% | 21% | 35% | 20% |
| K5 (Other female genital tract disorders) | 18% | 9% | 34% | 11% | 26% | 50% | 27% |
| N3 (Osteopathy/chondropathy/acquired musculoskeletal deformity) | 18% | 9% | 16% | 25% | 21% | 42% | 19% |
| S5 (Sprains and strains of joints and adjacent muscles) | 17% | 7% | 21% | 13% | 26% | 44% | 32% |
| SK (Traumatic complications and unspecified injuries) | 16% | 7% | 25% | 13% | 23% | 45% | 5% |
| S2 (Fracture of upper limb) | 15% | 11% | 26% | 18% | 15% | 23% | 18% |
| S3 (Fracture of lower limb) | 15% | 11% | 23% | 21% | 15% | 25% | 19% |
| B7 (Benign neoplasms) | 14% | 8% | 13% | 13% | 22% | 33% | 23% |
| G6 (Cerebrovascular disease) | 14% | 9% | 6% | 30% | 11% | 21% | 14% |
| J0 (Oral cavity, salivary glands and jaw diseases) | 14% | 5% | 22% | 12% | 20% | 44% | 16% |
| J6 (Liver, biliary, pancreas + gastrointestinal diseases NEC) | 14% | 7% | 28% | 23% | 13% | 30% | 14% |
| K2 (Male genital organ diseases) | 14% | 8% | 11% | 21% | 19% | 19% | 13% |
| C0 (Disorders of thyroid gland) | 13% | 8% | 9% | 18% | 14% | 24% | 16% |
| F3 (Peripheral nervous system disorders) | 13% | 5% | 11% | 17% | 16% | 33% | 16% |
| D0 (Deficiency anaemias) | 12% | 6% | 10% | 25% | 11% | 28% | 13% |
| F2 (Other central nervous system disorders) | 11% | 7% | 22% | 10% | 12% | 24% | 13% |
| K0 (Nephritis, nephrosis and nephrotic syndrome) | 11% | 6% | 8% | 27% | 8% | 19% | 8% |
| B3 (Malig neop of bone, connective tissue, skin and breast) | 10% | 6% | 2% | 14% | 15% | 19% | 12% |

| chapter | cohort | 1 | 2 | 3 | 4 | 5 | 6 |
|---|---|---|---|---|---|---|---|
| BB ([M]Morphology of neoplasms) | 10% | 6% | 5% | 13% | 14% | 20% | 10% |
| G7 (Arterial, arteriole and capillary disease) | 10% | 5% | 6% | 23% | 10% | 21% | 11% |
| J4 (Noninfective enteritis and colitis) | 9% | 4% | 14% | 9% | 11% | 25% | 8% |
| TJ (Drugs and other substances-adverse effects in theraputic use) | 9% | 3% | 5% | 16% | 9% | 27% | 22% |
| C2 (Nutritional deficiencies) | 8% | 4% | 8% | 16% | 6% | 18% | 5% |
| D2 (Aplastic and other anaemias) | 8% | 3% | 6% | 19% | 7% | 22% | 8% |
| K3 (Disorders of breast) | 8% | 3% | 13% | 5% | 11% | 28% | 14% |
| SD (Superficial injury) | 8% | 3% | 10% | 7% | 12% | 25% | 11% |
| SE (Contusion (bruise) with intact skin) | 8% | 3% | 9% | 9% | 11% | 29% | 8% |
| F1 (Hereditary and degenerative diseases of the CNS) | 7% | 5% | 6% | 10% | 8% | 16% | 6% |
| SN (Other and unspecified external effect causes) | 7% | 3% | 8% | 7% | 9% | 20% | 8% |
| SP (Surgical and medical care complications NEC) | 7% | 2% | 7% | 10% | 9% | 25% | 6% |
| H5 (Other respiratory system diseases) | 6% | 3% | 8% | 14% | 5% | 14% | 5% |
| S6 (Intracranial injury excluding those with skull fracture) | 6% | 3% | 18% | 6% | 6% | 13% | 7% |
| S8 (Open wound of head, neck and trunk) | 6% | 2% | 11% | 5% | 7% | 14% | 3% |
| B4 (Malignant neoplasm of genitourinary organ) | 5% | 4% | 2% | 8% | 7% | 6% | 6% |
| E1 (Non-organic psychoses) | 5% | 3% | 24% | 3% | 4% | 11% | 5% |
| R2 ([D]Cause of morbidity and mortality unsure and ill-defined) | 5% | 2% | 4% | 7% | 5% | 12% | 3% |
| T1 (Motor vehicle traffic accidents (MVTA)) | 5% | 3% | 11% | 4% | 6% | 9% | 5% |
| TE (Accidents due to natural and environmental factors) | 5% | 2% | 6% | 4% | 8% | 15% | 4% |
| G4 (Pulmonary circulation diseases) | 4% | 2% | 3% | 8% | 3% | 7% | 4% |
| K4 (Female pelvic inflammatory diseases) | 4% | 1% | 11% | 2% | 5% | 18% | 4% |
| L0 (Pregnancy with abortive outcome) | 4% | 3% | 16% | 2% | 4% | 8% | 5% |
| Ny (Musculoskeletal or connective tissue diseases OS) | 4% | 2% | 3% | 5% | 4% | 10% | 3% |
| S1 (Fracture of neck and trunk) | 4% | 2% | 7% | 6% | 4% | 9% | 5% |
| SL (Poisoning) | 4% | 2% | 39% | 2% | 1% | 4% | 3% |
| U6 ([X]Complications of medical and surgical care) | 4% | 1% | 1% | 10% | 3% | 10% | 2% |
| B1 (Malignant neoplasm of digestive organs and peritoneum) | 3% | 2% | 1% | 4% | 4% | 4% | 3% |
| B8 (Carcinoma in situ) | 3% | 2% | 1% | 4% | 4% | 6% | 4% |
| E0 (Organic psychotic conditions) | 3% | 2% | 13% | 4% | 2% | 5% | 2% |
| Fy (Other specified diseases of nervous system or sense organ) | 3% | 1% | 6% | 5% | 3% | 8% | 7% |
| S4 (Dislocations and subluxations) | 3% | 2% | 3% | 3% | 3% | 5% | 3% |
| S9 (Open wound of upper limb) | 3% | 1% | 6% | 2% | 3% | 7% | 1% |
| B6 (Malignant neoplasm of lymphatic and haemopoietic tissue) | 2% | 1% | 1% | 2% | 3% | 3% | 2% |
| D4 (White blood cell and other blood disorders) | 2% | 1% | 4% | 3% | 2% | 5% | 2% |
| H0 (Acute respiratory infections) | 2% | 1% | 1% | 6% | 1% | 4% | 1% |
| L2 (Risk factors in pregnancy) | 2% | 2% | 6% | 1% | 2% | 3% | 5% |
| L3 (Complications occurring during labour and delivery) | 2% | 1% | 7% | 1% | 2% | 3% | 3% |
| S0 (Fracture of skull) | 2% | 1% | 8% | 1% | 1% | 2% | 1% |
| SA (Open wound of lower limb) | 2% | 1% | 2% | 3% | 3% | 7% | 2% |
| SH (Burns) | 2% | 1% | 6% | 2% | 3% | 6% | 2% |
| U3 ([X]Assault) | 2% | 1% | 16% | 1% | 1% | 3% | 1% |
| A3 (Other bacterial diseases) | 1% | 0% | 1% | 2% | 0% | 1% | 0% |

| chapter | cohort | 1 | 2 | 3 | 4 | 5 | 6 |
|---|---|---|---|---|---|---|---|
| B2 (Malig neop of respiratory tract and intrathoracic organs) | 1% | 1% | 1% | 2% | 1% | 1% | 1% |
| B5 (Malignant neoplasm of other and unspecified sites) | 1% | 1% | 0% | 1% | 1% | 1% | 3% |
| B9 (Neoplasms of uncertain behaviour) | 1% | 1% | 0% | 2% | 1% | 2% | 1% |
| D3 (Clotting and bleeding disorders) | 1% | 1% | 2% | 2% | 2% | 4% | 1% |
| G. (Circulatory system diseases) | 1% | 0% | 0% | 0% | 0% | 0% | 55% |
| G1 (Chronic rheumatic heart disease) | 1% | 0% | 0% | 4% | 1% | 3% | 1% |
| H2 (Pneumonia and influenza) | 1% | 0% | 1% | 3% | 0% | 2% | 1% |
| H4 (Lung disease due to external agents) | 1% | 1% | 1% | 2% | 0% | 1% | 1% |
| J. (Digestive system diseases) | 1% | 0% | 0% | 0% | 0% | 0% | 67% |
| J2 (Appendicitis and other disorders of the appendix) | 1% | 0% | 1% | 1% | 1% | 1% | 1% |
| L1 (Pregnancy complications) | 1% | 1% | 6% | 0% | 1% | 3% | 1% |
| M. (Skin and subcutaneous tissue diseases) | 1% | 0% | 0% | 0% | 0% | 0% | 68% |
| Mz (Skin and subcutaneous tissue disease NOS) | 1% | 0% | 1% | 1% | 2% | 7% | 5% |
| N. (Musculoskeletal and connective tissue diseases) | 1% | 0% | 0% | 0% | 0% | 0% | 51% |
| Nz (Musculoskeletal and connective tissue diseases NOS) | 1% | 0% | 0% | 0% | 1% | 3% | 11% |
| PD (Urinary system congenital anomalies) | 1% | 0% | 1% | 1% | 1% | 1% | 1% |

Supplementary table 10: Code characteristics of cohort and clusters at Read code chapter level 3 (Percentage with the code in the cohort/cluster). Presenting 100 of 736 codes total.

| chapter | cohort | 1 | 2 | 3 | 4 | 5 | 6 |
|---|---|---|---|---|---|---|---|
| N24 (Other soft tissue disorders) | 54% | 30% | 51% | 69% | 77% | 96% | 57% |
| G20 (Essential hypertension) | 45% | 36% | 19% | 63% | 50% | 60% | 44% |
| N09 (Other and unspecified joint disorders) | 36% | 16% | 30% | 42% | 55% | 82% | 47% |
| N14 (Other and unspecified back disorders) | 35% | 17% | 36% | 39% | 50% | 78% | 42% |
| R00 ([D]General symptoms) | 33% | 14% | 41% | 40% | 46% | 80% | 35% |
| N05 (Osteoarthritis and allied disorders) | 31% | 16% | 12% | 44% | 43% | 65% | 37% |
| N21 (Peripheral enthesopathies and allied syndromes) | 28% | 13% | 21% | 33% | 42% | 63% | 41% |
| R02 ([D]Symptoms affecting skin and other integumentary tissue) | 27% | 11% | 24% | 32% | 39% | 68% | 33% |
| C10 (Diabetes mellitus) | 26% | 21% | 19% | 44% | 23% | 36% | 27% |
| F50 (Disorders of external ear) | 23% | 11% | 20% | 24% | 37% | 53% | 25% |
| G2. (Hypertensive disease) | 23% | 16% | 8% | 33% | 28% | 38% | 25% |
| F4C (Disorders of conjunctiva) | 22% | 10% | 18% | 22% | 32% | 55% | 26% |
| M03 (Other cellulitis and abscess) | 21% | 10% | 22% | 30% | 24% | 46% | 22% |
| N13 (Other cervical disorders) | 20% | 7% | 18% | 21% | 31% | 57% | 21% |
| R06 ([D]Respiratory system and chest symptoms) | 20% | 7% | 22% | 30% | 24% | 58% | 33% |
| TC. (Accidental falls) | 20% | 9% | 17% | 29% | 24% | 49% | 16% |
| E20 (Neurotic disorders) | 19% | 10% | 51% | 15% | 21% | 44% | 29% |
| R09 ([D]Other abdominal and pelvic symptoms) | 19% | 7% | 26% | 20% | 25% | 54% | 23% |
| C32 (Disorders of lipoid metabolism) | 18% | 11% | 10% | 31% | 20% | 33% | 14% |
| M12 (Contact dermatitis and other eczemas) | 18% | 8% | 18% | 18% | 27% | 47% | 14% |
| G57 (Cardiac dysrhythmias) | 17% | 10% | 5% | 41% | 15% | 29% | 18% |
| J10 (Diseases of oesophagus) | 17% | 7% | 19% | 24% | 21% | 46% | 18% |
| M22 (Other dermatoses) | 17% | 7% | 4% | 21% | 28% | 36% | 26% |
| J16 (Disorders of stomach function) | 15% | 6% | 17% | 18% | 21% | 44% | 24% |
| M2y (Other specified diseases of skin or subcutaneous tissue) | 15% | 7% | 9% | 19% | 24% | 36% | 16% |
| SK1 (Other specified injury) | 15% | 6% | 23% | 12% | 22% | 42% | 4% |
| F42 (Other retinal disorders) | 14% | 8% | 5% | 29% | 13% | 25% | 14% |
| F46 (Cataract) | 14% | 7% | 3% | 29% | 16% | 29% | 17% |
| H33 (Asthma) | 14% | 9% | 26% | 17% | 14% | 27% | 15% |
| Eu3 ([X]Mood - affective disorders) | 13% | 8% | 44% | 12% | 12% | 22% | 14% |
| F58 (Other ear disorders) | 13% | 5% | 12% | 12% | 21% | 37% | 15% |
| F59 (Hearing loss) | 13% | 7% | 6% | 19% | 18% | 26% | 14% |
| J52 (Functional gastrointestinal tract disorders NEC) | 13% | 5% | 16% | 14% | 17% | 36% | 20% |
| J57 (Other disorders of intestine) | 13% | 5% | 14% | 17% | 16% | 35% | 15% |
| K15 (Cystitis) | 13% | 6% | 13% | 13% | 18% | 37% | 13% |
| M18 (Pruritus and related conditions) | 13% | 3% | 10% | 14% | 19% | 45% | 14% |
| G83 (Varicose veins of the legs) | 12% | 6% | 6% | 17% | 17% | 29% | 18% |
| G84 (Haemorrhoids) | 12% | 5% | 13% | 13% | 17% | 31% | 15% |
| N22 (Other disorders of the synovium, tendon and bursa) | 12% | 5% | 8% | 12% | 19% | 34% | 16% |
| C04 (Acquired hypothyroidism) | 11% | 7% | 7% | 16% | 12% | 20% | 13% |
| C34 (Gout) | 11% | 6% | 4% | 22% | 12% | 17% | 9% |

| chapter | cohort | 1 | 2 | 3 | 4 | 5 | 6 |
|---|---|---|---|---|---|---|---|
| F4D (Inflammation of eyelids) | 11% | 4% | 9% | 10% | 16% | 29% | 12% |
| G3. (Ischaemic heart disease) | 11% | 5% | 3% | 34% | 7% | 20% | 10% |
| J51 (Diverticula of intestine) | 11% | 4% | 4% | 19% | 12% | 28% | 15% |
| K19 (Other urethral and urinary tract disorders) | 11% | 5% | 6% | 18% | 13% | 24% | 9% |
| M07 (Other local infections of skin and subcutaneous tissue) | 11% | 4% | 11% | 11% | 16% | 34% | 4% |
| M11 (Atopic dermatitis and related conditions) | 11% | 6% | 11% | 12% | 15% | 23% | 26% |
| N11 (Spondylosis and allied disorders) | 11% | 4% | 5% | 15% | 15% | 33% | 12% |
| N33 (Other bone and cartilage disorders) | 11% | 5% | 8% | 17% | 12% | 30% | 11% |
| R04 ([D]Head and neck symptoms) | 11% | 4% | 12% | 12% | 14% | 34% | 16% |
| D00 (Iron deficiency anaemias) | 10% | 5% | 9% | 22% | 9% | 25% | 11% |
| E22 (Sexual deviations or disorders) | 10% | 6% | 10% | 14% | 13% | 13% | 15% |
| E2B (Depressive disorder NEC) | 10% | 4% | 28% | 7% | 11% | 32% | 15% |
| F56 (Vertiginous syndromes, other disorders of vestibular system) | 10% | 4% | 5% | 10% | 15% | 28% | 13% |
| G33 (Angina pectoris) | 10% | 4% | 3% | 28% | 7% | 21% | 13% |
| J34 (Diaphragmatic hernia) | 10% | 4% | 8% | 17% | 10% | 24% | 10% |
| M15 (Erythematous conditions) | 10% | 3% | 8% | 10% | 14% | 30% | 10% |
| G30 (Acute myocardial infarction) | 9% | 5% | 3% | 27% | 5% | 12% | 7% |
| G58 (Heart failure) | 9% | 4% | 2% | 32% | 3% | 15% | 8% |
| G80 (Phlebitis and thrombophlebitis) | 9% | 4% | 7% | 12% | 11% | 23% | 10% |
| M0. (Skin and subcutaneous tissue infections) | 9% | 4% | 11% | 12% | 11% | 22% | 6% |
| B76 (Benign neoplasm of skin) | 8% | 4% | 8% | 6% | 13% | 18% | 13% |
| C36 (Disorders of fluid, electrolyte and acid-base balance) | 8% | 3% | 8% | 16% | 6% | 22% | 7% |
| D21 (Other and unspecified anaemias) | 8% | 3% | 5% | 19% | 7% | 22% | 8% |
| J15 (Gastritis and duodenitis) | 8% | 3% | 14% | 14% | 8% | 24% | 9% |
| K5A (Menopausal and postmenopausal disorders) | 8% | 3% | 8% | 5% | 13% | 33% | 12% |
| M23 (Diseases of nail) | 8% | 3% | 7% | 8% | 11% | 22% | 13% |
| N06 (Other and unspecified arthropathies) | 8% | 3% | 4% | 11% | 11% | 25% | 5% |
| R08 ([D]Urinary system symptoms) | 8% | 3% | 6% | 12% | 10% | 23% | 14% |
| R1y ([D]Other specified nonspecific abnormal findings) | 8% | 4% | 4% | 9% | 11% | 18% | 5% |
| B33 (Other malignant neoplasm of skin) | 7% | 4% | 1% | 12% | 11% | 13% | 8% |
| C11 (Other disorders of pancreatic internal secretion) | 7% | 4% | 6% | 13% | 7% | 13% | 7% |
| C38 (Obesity and other hyperalimentation) | 7% | 3% | 9% | 9% | 8% | 17% | 10% |
| F45 (Glaucoma) | 7% | 5% | 2% | 11% | 8% | 11% | 7% |
| F4K (Other eye disorders) | 7% | 2% | 4% | 8% | 11% | 26% | 7% |
| G73 (Other peripheral vascular disease) | 7% | 3% | 4% | 16% | 6% | 13% | 9% |
| H3. (Chronic obstructive pulmonary disease) | 7% | 4% | 10% | 19% | 3% | 11% | 5% |
| K31 (Other breast disorders) | 7% | 2% | 12% | 5% | 9% | 27% | 7% |
| K59 (Menstruation disorders) | 7% | 3% | 22% | 2% | 8% | 17% | 10% |
| M26 (Sebaceous gland diseases) | 7% | 4% | 13% | 4% | 11% | 17% | 10% |
| M27 (Chronic skin ulcer) | 7% | 3% | 4% | 14% | 7% | 17% | 6% |
| S23 (Fracture of radius and ulna) | 7% | 5% | 9% | 8% | 7% | 13% | 9% |
| Eu0 ([X]Organic, including symptomatic, mental disorders) | 6% | 5% | 2% | 12% | 5% | 8% | 2% |
| F26 (Migraine) | 6% | 3% | 12% | 4% | 8% | 18% | 9% |

| chapter | cohort | 1 | 2 | 3 | 4 | 5 | 6 |
|---|---|---|---|---|---|---|---|
| F34 (Mononeuritis of upper limb and mononeuritis multiplex) | 6% | 3% | 5% | 8% | 9% | 17% | 7% |
| F51 (Nonsuppurative otitis media + eustachian tube disorders) | 6% | 3% | 8% | 4% | 11% | 19% | 7% |
| F52 (Suppurative and unspecified otitis media) | 6% | 3% | 11% | 4% | 10% | 18% | 3% |
| G65 (Transient cerebral ischaemia) | 6% | 3% | 2% | 14% | 5% | 11% | 7% |
| J08 (Oral soft tissue disease) | 6% | 2% | 7% | 5% | 8% | 22% | 7% |
| J30 (Inguinal hernia) | 6% | 4% | 3% | 10% | 8% | 8% | 7% |
| J43 (Other non-infective inflammatory gastroenteritis and colitis) | 6% | 3% | 11% | 6% | 8% | 20% | 5% |
| K04 (Acute renal failure) | 6% | 3% | 6% | 15% | 4% | 10% | 4% |
| M10 (Erythematosquamous dermatosis) | 6% | 2% | 6% | 6% | 9% | 15% | 11% |
| M16 (Psoriasis and similar disorders) | 6% | 4% | 6% | 6% | 8% | 11% | 7% |
| M21 (Other atrophic and hypertrophic conditions of skin) | 6% | 2% | 4% | 6% | 10% | 19% | 7% |
| M24 (Hair and hair follicle diseases) | 6% | 2% | 10% | 4% | 8% | 18% | 9% |
| M2z (Other skin and subcutaneous tissue disease NOS) | 6% | 2% | 3% | 7% | 9% | 15% | 2% |
| N23 (Muscle, ligament and fascia disorders) | 6% | 2% | 6% | 6% | 8% | 15% | 8% |
| R11 ([D]Nonspecific urine findings) | 6% | 3% | 2% | 14% | 5% | 10% | 7% |
| S64 (Intracranial injury NOS) | 6% | 3% | 17% | 6% | 6% | 12% | 6% |
